# Divergent T Cell Phenotypes Define Pediatric Crohn’s Disease and Ulcerative Colitis

**DOI:** 10.1101/2025.10.16.25336112

**Authors:** Leonard Nettey, Hengqi Betty Zheng, Joseph C. Devlin, Julie E. Horowitz, Sara Rosenbaum, Nathalie Fiaschi, Wei Keat Lim, Kyle Kimler, Christina Adler, Annabelle Lee, Min Ni, Yi Wei, Peter J. Ehmann, Shane E. McCarthy, Manuel A.R. Ferreira, William Galbavy, Eli Stahl, Sokol Haxhinasto, Sandra Coetzee, Yoko Yabe, Michael Dobosz, Paula Keskula, Jillian Zavistaski, Alexandre Albanese, Zoë Steier, Joshua de Sousa Casal, Veronika Niederlova, Lusine Ambartsumyan, Ghassan Wahbeh, David L. Suskind, Vanessa Mitsialis, Sara Hamon, Stephen M. Carpenter, Jennifer D. Hamilton, Alex K. Shalek, Scott B. Snapper, Matthew A. Sleeman, George D. Kalliolias, Andrea T. Hooper, Jose Ordovas-Montanes, Leslie S. Kean

## Abstract

Pediatric Inflammatory Bowel Disease (IBD) remains challenging to treat and difficult to prognosticate. Although multiple immune cell types coordinate pathology in both Crohn’s disease (CD) and ulcerative colitis (UC), specifying which cell types and cell states portend better or worse response to major IBD treatment strategies, including anti-TNF therapies (the only FDA-approved therapy for pediatric IBD), remains challenging. Here, we present the results of the PREDICT study, which enrolled 79 treatment-naïve pediatric patients at the time of diagnostic endoscopy, enabling a comprehensive transcriptomic, histologic, and serologic analysis of 40 CD patients and 16 UC patients, as well as 23 patients with functional gastrointestinal disorders (FGID) who served as pediatric non-inflamed controls. Leveraging these data, we performed a comprehensive analysis of colonic immunology in each of these clinical conditions. Our results indicate that, within the complex landscape of immune pathology in pediatric CD and UC, there is a coordinated shift across the T_H_1-to-T_H_17 immune activation continuum among T cells that is pertinent to anti-TNF response. For CD, this shift defines partial response to anti-TNF treatment. For UC, the landscape is more complex, with both T_H_17 and TFH biology defining disease, and pre-treatment T_H_17 biology contributing to anti-TNF treatment resistance. Related to this, polyreactive TCR phenotypes within UC T_FH_ cells are correlated with both germinal center activity and the frequency of IgG1 plasma cells, yet opposed to the T_H_17 signatures associated with anti-TNF nonresponse. The elucidation of these distinct mechanisms of T cell-dependent disease pathology, treatment response, and TCR polyreactivity suggests a model in which sustained T_H_17 signaling in CD and baseline T_H_17 signaling in UC are associated with disease pathogenesis, forming a basis for a generalized understanding of IBD pathogenesis and underscoring the need for endotype-specific approaches to IBD therapy.

## INTRODUCTION

Inflammatory bowel disease (IBD), for which Crohn’s disease (CD) and ulcerative colitis (UC) are the most common subtypes, encompasses a group of debilitating inflammatory gastrointestinal conditions with increasing incidence^1^. Although pediatric and adult disease have many clinical and biologic commonalities, pediatric IBD can be particularly devastating, given that this disease is often treatment refractory, more predisposed to rapid progression^2^, and requires effective treatment for many decades. Recent efforts have catalogued immunologic associations with treatment resistance in biologic-, but not treatment-naïve, adults; however, no study has yet defined the comparative landscape of these diseases at diagnosis (naïve to all treatments), nor provided a multiomic map of these diseases in pediatric patients^3^. Understanding the cellular and molecular immunopathologic networks causing pediatric disease, elucidating the mechanisms of treatment failure in these young patients, and identifying novel avenues for therapeutic intervention^4^ remain major unmet clinical needs.

Acknowledging the central role that T cells play in both CD and UC, multiple therapies targeting T cell trafficking, IL-12/IL-23, and lymphocyte activation are in use. However, anti-TNF monoclonal antibodies remain first-line biologic treatment for both CD and UC^5,6^, and the only FDA-approved biologic therapies for pediatric IBD. The success of anti-TNF agents is highly variable, and there remains no diagnostic paradigm for prioritizing other agents due to limited understanding of TNF-resistant cell states in CD or UC. Such functional knowledge is urgently required, given that differing mechanisms of IBD pathology may be better treated by biologic therapies targeted towards disease endotypes.

To address these knowledge gaps, we created a prospective natural history study, the Precision Diagnostics in Inflammatory Bowel Disease, Cellular Therapy, and Transplantation study (“PREDICT”, NCT03369353), which enrolled and evaluated a cohort of treatment-naïve pediatric patients (≥ 6 years old) with GI distress requiring endoscopy (n = 79), and who were diagnosed with one of four conditions: non-inflammatory ‘Functional GI Disease’ (FGID), ileal-only CD (iCD), colon-involving CD (cCD), or UC. Key to the design of this study was the enrollment of patients prior to diagnostic endoscopy and any treatment, such that the distinctive immunobiology of these conditions could be determined, unaffected by concomitant immunosuppressive treatments. PREDICT combines detailed clinical evaluation and follow-up with multiomic biologic evaluation including 5’-scRNA-seq/scTCR-seq, multiplexed immunohistochemistry (IHC), and human proteome array-based autoantibody detection. This study enabled us to uncover distinct cellular disease mechanisms that drive pathology and treatment resistance within pediatric CD and UC, providing an evidence-based foundation for future evaluations of molecular disease-targeted therapies in these patients.

## RESULTS

### PREDICT: Clinical Study Enrollment, Clinical Outcomes, and Follow-Up

PREDICT IBD study participants were enrolled from November 2017 to January 2022, during which time 23 FGID, 7 iCD, 33 cCD (1 with perianal-only disease, and 2 without successfully processed diagnostic scRNA-seq biopsies), and 16 UC patients were enrolled (**Fig. 1a**). In addition to the diagnostic biopsies, 4 iCD, 10 cCD, and 2 UC patients contributed follow-up biopsies that were processed for downstream analysis. At diagnosis, enrolled patients underwent clinical and laboratory evaluation, and both upper and lower endoscopy to evaluate disease and obtain biopsies. Following endoscopy, standard clinical and histologic features were used by the treating physicians to classify participants as FGID, iCD, cCD, or UC, and all treatment decisions were made by the treating physicans^7^. After their initial diagnosis, patients with IBD were followed clinically for up to 2 years. Patients with FGID were followed clinically only as needed. Of the FGID, iCD, cCD, and UC patients analyzed, the median age at diagnosis was 15.7, 17.6, 11.4, and 12.8, respectively (p = 0.013, ANOVA, **Supplemental Table 1**). Males represented 56.5, 71.4, 54.5, and 56.3% of patients with FGID, iCD, cCD, and UC, respectively (p = 0.88, chi-squared test, **Supplemental Table 1**).

**Fig. 1.**
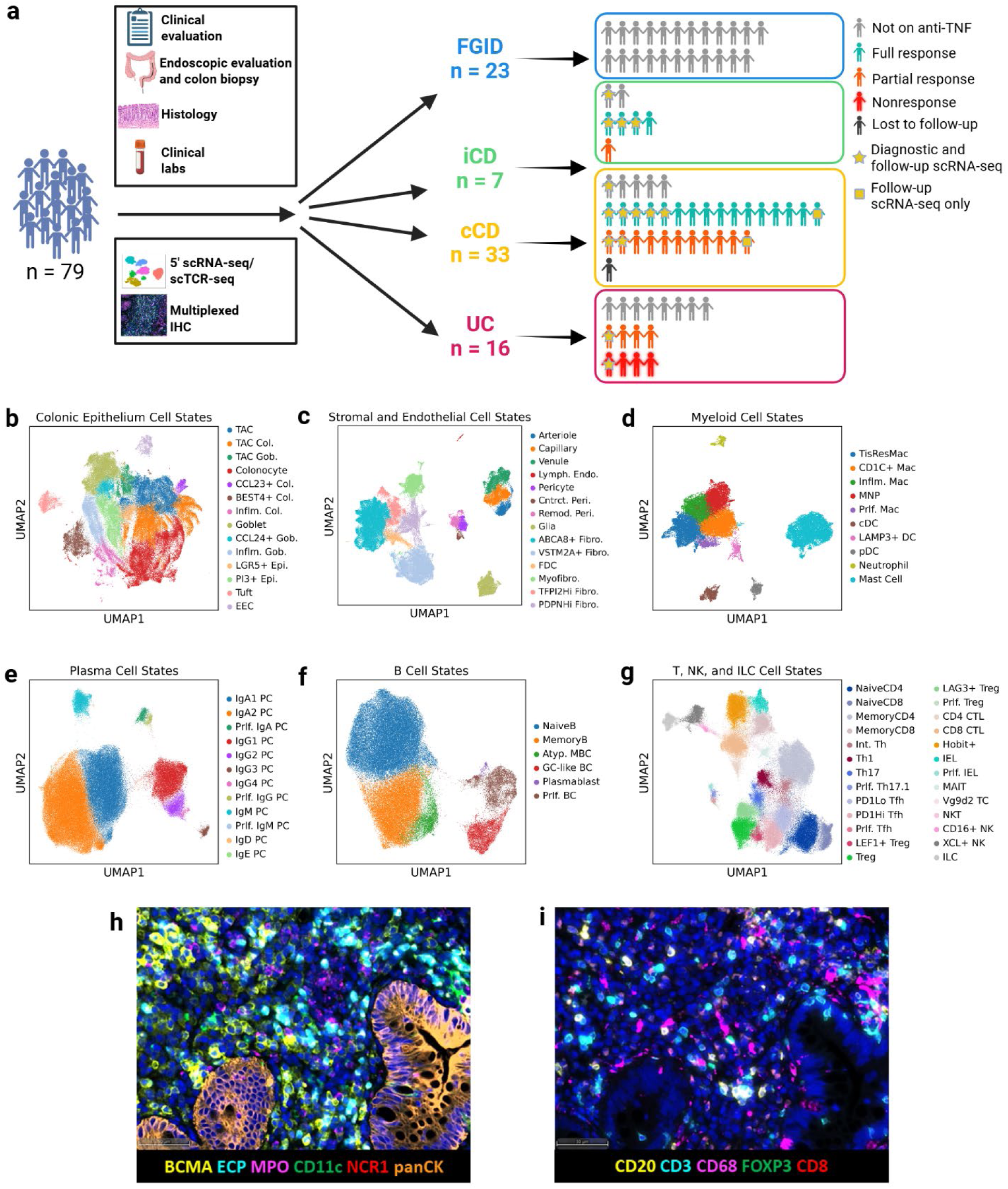
PREDICT Study Overview, Cell States, and Immunohistochemistry Panels. **a**, Outline of PREDICT data collection, specimen collection, and treatment response. During each endoscopy, colon biopsy and clinical metadata are obtained. Additional clinical metadata are obtained throughout the study’s 2 year follow-up period. Additional colon biopsies obtained if clinically indicated. Primary assays performed with colon biopsies include 5’ scRNA-seq and scTCR-seq of dissociated epithelium and lamina propria and *in situ* fluorescence microscopy. Visual representation of study participants with diagnostic colon biopsy successfully processed for scRNA-seq per disease category (FGID, n = 23; iCD, n = 7, cCD, n = 33; UC, n = 16; overall, n = 79). Participants with successfully processed diagnostic and follow-up biopsies are indicated by yellow star (FGID, n = 0; iCD, n = 4; cCD, n = 8; UC, n = 2). Participants with only follow-up biopsy successfully processed (i.e. without successfully processed diagnostic biopsy) indicated by yellow square (FGID, n = 0; iCD, n = 0; cCD, n = 2; UC, n = 0). Participant groups are colored by disease category. Blue, FGID. Green, iCD. Yellow, cCD. Red, UC. Participant images are colored by anti-TNF response category. Dark gray, lost to follow-up. Light gray, NOA. Cyan, FR. Orange, PR. Red with glow, NR. **b**, **c**, **d**, **e**, **f**, **g**, UMAPs of cell state annotations for each cell type: (**b**) colonic epithelium, (**c**) stromal and endothelial cells, (**d**) myeloid cells, (**e**) plasma cells, (**f**) B cells, and (**g**) T cells, natural killer cells, and innate lymphoid cells (T, NK, and ILCs). **h**, **i**, Representative micrographs of immunohistochemical panels of colon biopsies from PREDICT study participants (images shown are from colon of participant 001-047). Two immunophenotyping panels were used. Immunophenotyping panel 1 contains markers for plasma cells (BCMA), eosinophils (ECP), neutrophils (MPO), dendritic cells and monocytes (CD11c), and NK cells (NCR1). Immunophenotyping panel 2 contains markers for B cells (CD20), T cells (CD3), myeloid cells (CD68), Treg cells (FOXP3), and CD8 T cells (CD8). panCK was used as marker for epithelial cells. Scale bar denotes 50 μm. FGID, functional gastrointestinal disorder. iCD, ileal-only Crohn’s disease. cCD, colon-involving Crohn’s disease. UC, ulcerative colitis. TNF, tumor necrosis factor. NOA, not on anti-TNF. FR, full response. PR, partial response. NR, nonresponse. scRNA-seq, single-cell RNA sequencing. scTCR-seq, single-cell TCR sequencing. GEX, gene expression. TCR, T cell receptor. IHC, immunohistochemistry. UMAP, uniform manifold approximation and projection. CD, cluster of differentiation. BCMA, B cell maturation antigen. ECP, eosinophil cationic protein. MPO, myeloperoxidase. NCR1, natural cytotoxicity receptor 1. DAPI, 4′,6-diamidino-2-phenylindole. panCK, pan-cytokeratin. FOXP3, forkhead box P3.

The majority (70%) of total IBD patients were treated with anti-TNF, with both infliximab and adalimumab used, based on treating physician decision-making (**Fig. 1a, Supplemental Table 1**). Among patients with successfully processed diagnostic biopsies, 5/7 iCD, 25/31 cCD, and 8/16 UC patients received anti-TNF. For these patients, we defined anti-TNF response status at 2 years post-diagnostic endoscopy. Anti-TNF treatment response categorization [Full Response (FR), Partial Response (PR), Nonresponse (NR)] was adjudicated by two pediatric gastroenterologists (HBZ and SR) based on the following criteria: FR was defined as clinical symptom control and biochemical response (normalization of C-reactive protein (CRP) and erythrocyte sedimentation rate (ESR)) on maintenance anti-TNF therapy with dose adjustments only due to insufficient blood drug levels. PR was defined as improvement in clinical symptoms or CRP/ESR without full clinical symptom control or CRP/ESR normalization, with documented escalation of anti-TNF therapy or additional therapeutic agents added. NR was defined as lack of clinical and biochemical response to anti-TNF, using the criteria defined above for FR and PR. In PREDICT, 4/5 iCD, 15/25 cCD, and 0/8 UC patients achieved FR. 1/5 iCD, 10/25 cCD, and 4/8 UC patients achieved PR. 0/5 iCD, 0/25 cCD, and 4/8 UC patients were designated as NR (**Fig. 1a, Supplemental Table 1**). In addition, for each disease there were patients for whom the treating physician chose not to treat with anti-TNF agents (2/7 iCD, 6/31 cCD, and 8/16 UC). These patients are designated as “not on anti-TNF” (“NOA”), and were treated with a variety of other immunomodulators and antimetabolic agents, based on their treating physician’s choice.

### scRNA-seq and IHC analysis define parenchymal, mesenchymal, and immunological features in FGID, CD and UC

We performed cellular profiling on fresh (non-frozen) diagnostic and follow-up colon biopsies through linked 5’ scRNA-seq and scTCR-seq (**Fig. 1a**). During sample processing, biopsies were separated into lamina propria and epithelial fractions, and samples from each fraction were separately prepared for scRNA-seq^8,9^. The use of 5’ chemistry enabled us to simultaneously interrogate the gene expression profiles and TCR identities of the profiled T cells using scTCR-seq. Following quality control and cellular annotation, 650,501 high-quality gene expression transcriptomes from diagnostic and follow-up biopsies were further analyzed (**Supplemental Table 1**).

To annotate cells, we first identified major “cell types” through low-resolution clustering (see Methods)^10^. These cell types were analyzed agnostic to GI layer of origin and included colonic epithelium; stromal and endothelial cells; myeloid cells; plasma cells; B cells; and T cells, NK cells & innate lymphoid cells (**Fig. 1b-g**, **Supplemental Fig. 1a-g, 2a-f, Supplemental Table 2**). Each cell type was separately analyzed to identify additional levels of detail, which we termed “cell subtypes” and “cell states” (**Supplemental Fig. 1h**). We manually annotated a total of 82 cell states within 31 cell subtypes across the 6 cell types (**Supplemental Table 3**)^8,11,12^. Each final annotated cell state was represented by multiple samples, regardless of the number of cells comprising the cell state (**Supplemental Fig. 1a, Supplemental Fig. 2a-f**). In addition, IHC was performed on biopsies from 44 PREDICT participants (8 FGID, 6 iCD, 15 UC, 15 cCD). Two immunophenotyping panels were used, identifying B cells (CD20), T cells (CD3), myeloid cells (CD68), T_reg_ cells (FOXP3), CD8 T cells (CD8), plasma cells (BCMA), eosinophils (ECP), neutrophils (MPO), dendritic cells and monocytes (CD11c), NK cells (NCR1), and epithelial cells (panCK) (**Fig. 1h**, **i**).

### Transcriptomic and Histologic Analysis Reveal Numerous Cellular Distinctions Between FGID and IBD

To statistically characterize major differences in the cell state frequencies between FGID controls and IBD at diagnostic endoscopy, we used Milo, a tool that generates differential abundance statistics for scRNA-seq data by assigning cells to partially overlapping neighborhoods on a *k*-nearest neighbor graph^13^. Milo neighborhoods were created within each cell type to determine which cell states contributed most to the variation observed amongst FGID, cCD, and UC colon biopsies. When comparing IBD samples with colonic inflammation (both cCD and UC) to FGID controls, we found enrichment in multiple activated cell types, including inflamed epithelial cells, inflammatory fibroblasts, inflammatory macrophages, mononuclear phagocytes (MNPs), plasmacytoid dendritic cells (pDCs), neutrophils, and multiple activated and inflammatory T cell populations (**Fig. 2a**, **Supplemental Table 4**). These findings demonstrate a diverse mucosal inflammatory response in both cCD and UC. IHC analysis corroborated the overarching scRNA-seq findings, identifying increased densities of CD11c+ cells, MPO+ cells, total CD3+ cells, as well as CD3+CD8-FOXP3+ cells (T_regs_) in IBD samples (**Fig. 2b, Supplemental Table 5**).

**Fig. 2.**
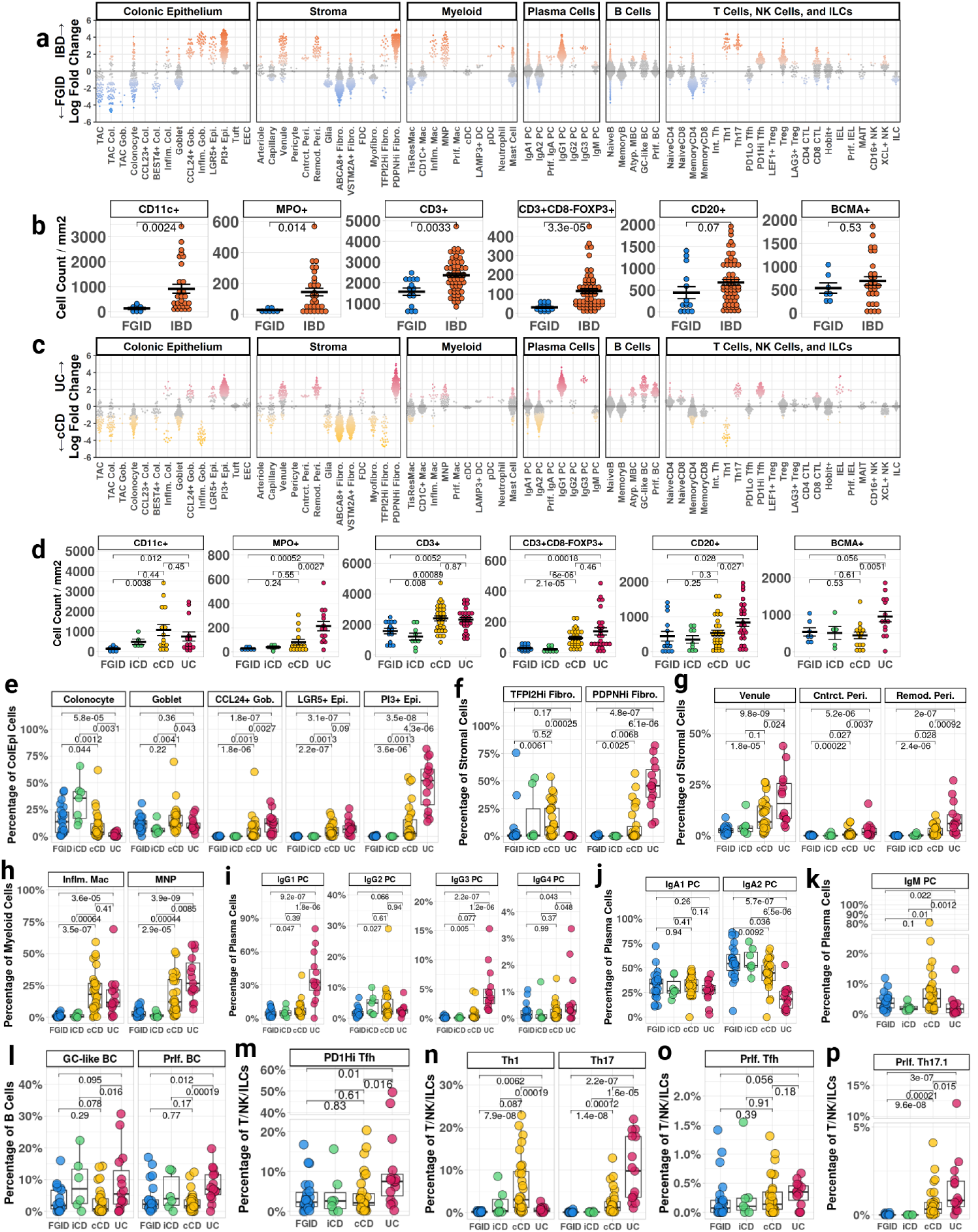
Transcriptomic and histochemical profiling highlights cellular shifts in cCD and UC. **a**, Milo-based differential abundance analysis performed within the six annotated cell types among all biopsies from diagnostic endoscopies, excluding biopsies with < 5 cells within the cell type after downsampling to equal cell numbers from FGID, cCD, and UC (FGID, n = 22; cCD, n = 31; UC, n = 16 for colonic epithelium. FGID, n = 23; cCD, n = 30; UC, n = 14 for stromal and endothelial cells. FGID, n = 23; cCD, n = 30; UC, n = 15 for myeloid cells and plasma cells. FGID, n = 21; cCD, n = 31; UC, n = 15 for B cells. FGID, n = 23; cCD, n = 31; UC, n = 15 for T cells, NK cells, and innate lymphoid cells). Each dot represents a grouping of cells with similar transcriptomic profiles (“neighborhood”). Identified neighborhoods were assigned to cell states based on the cell state with ≥ 70% representation within the neighborhood. Neighborhoods with no cell state occupying ≥ 70% of neighborhood were removed from analysis. Cell states not shown did not meet criteria in any neighborhood. Comparisons made between FGID and IBD (cCD and UC). Individual Milo-derived neighborhoods are colored by disease if their spatial FDR < 0.1. Blue = FGID. Orange = IBD. **b**, Cell densities of select markers in immunophenotyping panels, comparing FGID to IBD (cCD and UC). Panel with markers BCMA, ECP, MPO, CD11c, and NCR1 was stained only once per study participant (FGID, n = 7; cCD, n =15; UC, n = 14). Panel with markers CD20, CD3, CD68, FOXP3, and CD8 was stained 1-3 times per study participant (FGID, n = 8 participants, n = 14 samples; cCD, n =15 participants, n = 30 samples; UC, n = 14 participants, n = 27 samples). **c**, Milo-based differential abundance analysis performed as in (**a**), comparing cCD (yellow) and UC (red). **d**, Cell densities of select markers in immunophenotyping panels, performed as in (**c**), comparing FGID, iCD, cCD, and UC. **e-p**, scRNA-seq cell state frequency (relative to cell type) comparisons for non-inflamed and inflamed epithelial cell states (**e**), TFPI2Hi and PDPNHi fibroblasts (**f**), venules, contractile pericytes, and remodeling pericytes (**g**), inflammatory macrophages and mononuclear phagocytes (**h**), IgG plasma cells (**i**), IgA plasma cells (**j**), IgM plasma cells (**k**), germinal center-like and proliferating B cells (**l**), PD1Hi T_FH_ cells (**m**), T_H_1 and T_H_17 cells (**n**), proliferating T_FH_ cells (**o**), and proliferating T_H_17.1 cells (**p**). Only biopsies with at least 10 cells in the cell type were included for **e-p** cell state frequency analyses (FGID, n = 23; iCD, n = 7; cCD, n = 31; UC, n = 16 for colonic epithelilum. FGID, n = 23; iCD, n = 7; cCD, n = 30; UC, n = 14 for stromal and endothelial cells. FGID, n = 23; iCD, n = 7; cCD, n = 30; UC, n = 15 for myeloid cells and plasma cells. FGID, n = 21; iCD, n = 6; cCD, n = 31; UC, n = 15 for B cells. FGID, n = 23; iCD, n = 7; cCD, n = 31; UC, n = 15 for T cells, NK cells, and innate lymphoid cells). Mann-Whitney U test, two-tailed, used for all pairwise comparisons. Error bars denote standard error of the mean.

### Transcriptomic and Histologic Distinguishers of cCD and UC: Evidence for Increased Epithelial Damage in UC

While pediatric cCD and UC can be challenging to distinguish endoscopically and histologically^14^, Milo differential abundance identified multiple cell states that could clearly distinguish these two IBD entities (**Fig. 2c**). Compared to UC, cCD biopsies were enriched in epithelial cell states suggestive of retained epithelial function, including non-inflamed absorptive colonocytes and goblet cells (**Fig. 2c**). In contrast, most epithelial cell states were depleted in diagnostic samples from UC patients, and the cells that remained in UC were skewed towards *LGR5*+ epithelial stem cells, *CCL24*-expressing goblet cells, and an epithelial population with expression of antimicrobial and T_H_17-promoting genes (*PI3* and *SAA1*, respectively), previously demonstrated to be induced by proinflammatory cytokines, including TNF, IL-1β, and IL-6 (**Fig. 2c, Supplemental Fig. 2a**)^15–19^. These cell states are consistent with characteristic UC histopathology findings that reveal regenerating stem cells in the setting of inflamed epithelium lacking normally functioning absorptive and secretory cells^20^.

Consistent with the epithelial damage in UC, the fibroblasts that support differentiating epithelium (annotated as *ABCA8*+ and *VSTM2A*+ fibroblasts) were generally absent from UC biopsies, but present in cCD biopsies (**Fig. 2c**). We also identified inflammatory fibroblast populations that were nearly exclusive to either cCD (“TFPI2Hi fibroblasts”) or UC (“PDPNHi fibroblasts”) (**Fig. 2c**). TFPI2Hi fibroblasts exhibited greater expression of chemokines *CCL13* and *CXCL10*, both of which are induced by IFNγ signaling (**Supplemental Fig. 2b**)^21–23^. In contrast, PDPNHi fibroblasts were enriched for genes involved in extracellular matrix organization and vascular remodeling, as have been described in adult UC (**Supplemental Fig. 2b**)^12^. In line with increased vascular remodeling, UC biopsies demonstrated an enrichment in the venule cell state and pericytes involved in capillary constriction and vascular remodeling (**Fig. 2c**)^11,24^.

### Transcriptomic and Histologic Distinguishers of cCD and UC: Evidence for Increased Inflammatory Myeloid Activity and Enhanced Germinal Center Activity in UC

While enrichments in inflammatory myeloid populations were often shared between both cCD and UC biopsies compared to FGID (with notable elevations in MNPs, inflammatory macrophages, and pDCs, **Fig. 2a**), UC biopsies were more skewed towards these monocyte, macrophage, and dendritic cell inflammatory states compared to cCD (**Fig. 2c**). IHC analysis also revealed a marked increase in MPO+ cells in UC, indicating neutrophilia that was not found in cCD samples (**Fig. 2d**). Plasma cells were significantly shifted towards IgG_1_ and IgG_3_ in UC (**Fig. 2c**), as has been previously reported^25–27^. We also observed that, concomitant with the increase in IgG, there was a coordinated decrease in both IgM and IgA plasma cells among UC biopsies, with the latter dominated by a reduction in IgA_2_, rather than all IgA cells (**Fig. 2c**). Potentially connected to the shifts in the immunoglobulin balance, UC could be distinguished from cCD by the presence of proliferating B cells and germinal center-like B cells, which were both increased in UC versus cCD (**Fig. 2c**). UC samples were also enriched for PD1Hi T_FH_ cells, which are likely to be germinal center T_FH_ cells^28,29^ (**Fig. 2c)**. The combination of increases in these cell state frequencies suggested elevated germinal center activity in colonic mucosa from UC patients compared to both FGID and cCD, which likely contributed to the observed plasma cell shifts. These scRNA-seq findings were corroborated by IHC analysis which demonstrated increased CD20+ (B cells) and BCMA+ (plasma cells) cell density in UC relative to cCD (**Fig. 2d**).

### Transcriptomic and Histologic Distinguishers of cCD and UC: Evidence for skewing towards T_H_1 immunity for CD, and T_H_17 immunity for UC at the time of diagnosis

T cells play a major role in IBD, coordinating interactions with immune, stromal, and parenchymal cells that contribute to inflammation, with known instructive roles for T_H_1 and T_H_17 cells in CD and UC^3,8,9,30–32^. We found that both T_H_1 and T_H_17 cell states were enriched in cCD and UC relative to FGID (**Fig. 2a**). Moreover, testing for differences between cCD and UC highlighted a notable shift towards T_H_1 cells in cCD and towards T_H_17 cells in UC (**Fig. 2c**). Milo-based differential abundance analysis was corroborated with per-biopsy comparisons of FGID, iCD, cCD, and UC cell state frequencies (**Fig. 2e-n**, **Supplemental Table 5**). Given the well-established connections between inflammatory T cell signaling and T cell proliferation^33^, we explicitly interrogated proliferating T cell states in CD versus UC. Because proliferating T cell states did not comprise enough cellular neighborhoods for Milo analysis (see Methods), this analysis was limited to per-biopsy comparisons. Our findings were consistent with the observation described above, of more PD1Hi T_FH_ in UC versus CD (**Fig. 2c**), with a trend towards more proliferating T_FH_ cells also observed in UC (**Fig. 2o**). Because proliferating T cells often express transcriptomic hallmarks of both T_H_1 and T_H_17 cells^34,35^, we defined a proliferating T cell state with features of both (including *CXCR3*, *IFNG*, *IL17A*, and *GNLY*, termed “Prlf. T_H_17.1”). Interrogation of Prlf. T_H_17.1 cells revealed that they were enriched in both cCD and UC compared to FGID, and of the two disease states, most enriched in patients with UC (**Fig. 2p, Supplemental Fig. 2f**).

### T_H_ cells are associated with increased T cell activation and exhaustion in UC

Having identified increased T_H_1 frequency in cCD and increased T_H_17 frequency in UC, we investigated whether detailed analysis of these T cell populations could further elucidate distinct signatures in each IBD subtype. Given the plasticity of helper T cells and their divergent association with cCD and UC, we performed differential gene expression comparing UC and cCD within the “T helper” cell subtype, which includes T_H_1, T_H_17, and Prlf. T_H_17.1 cells. This analysis identified genes enriched in UC versus cCD related to T cell activation, co-stimulation, or exhaustion, including *TOX*, *TNFRSF4* (OX40), *TNFRSF18* (GITR), *LAG3*, and *CD247* (CD3ζ) (**Supplemental Fig. 3a**, **Supplemental Table 6**). We further delineated these DE genes within the T_H_1 and T_H_17 cell states. We found that these genes, as well as *HAVCR2* (TIM-3), were increased in both UC T_H_17 and UC T_H_1 cells, confirming that their enrichment in the T_H_ cell subtype was not solely due to differences in cell state abundance (**Supplemental Fig. 3b**, **Supplemental Table 7**).

Next, we investigated the ambiguity of the Prlf. T_H_17.1 cell state by analyzing the expression of T_H_1- and T_H_17-identifying genes within the Prlf. T_H_17.1 cell state, as well as within T_H_1, and T_H_17 cell states. Even amongst cells annotated as T_H_1, we found increased expression of T_H_1-associated genes (*IFNG*, *TNF*, *ANXA1*, *CXCR3*, *GZMK*) in cCD compared to UC (**Supplemental Fig. 3c**). Importantly, *TNF*, *ANXA1*, *CXCR3*, and *GZMK* were also increased in cCD Prlf. T_H_17.1 cells, suggesting that T_H_1 cells may be the primary cell state of origin in cCD Prlf. T_H_17.1 cells (**Supplemental Fig. 3c**). Conversely, genes *IL17A*, *IL17F*, *IL26*, and *GNLY* were increased in UC Prlf. T_H_17.1 cells relative to cCD, suggesting an association with the T_H_17 cell state (**Supplemental Fig. 3d**), and distinguishing the origin of these cells in the two IBD endotypes. Notably, we observed increased *IL17A* expression in UC T_H_1 cells. In the context of increased T cell activation and exhaustion in UC, these data suggest that UC T_H_1 cells may actually be ex-T_H_17 cells, known to take on a T_H_1-like phenotype under prolonged inflammatory conditions^36^.

### Clonal overlap within T_H_ cell states highlights enrichment for *CD8A+* T_H_17 cells in UC

We took advantage of the 5’ scRNA-seq chemistry to simultaneously track linked TCR sequences, to determine the extent to which T_H_1 or T_H_17 clones shared TCRs with proliferating T_H_17.1 cells in cCD or UC (**Supplemental Fig. 3e-g**, **Supplemental Table 8**). We found that Prlf. T_H_17.1 clones came from both T_H_1 and T_H_17 cell states in cCD, with a trend towards increased representation of T_H_1 clones (**Supplemental Fig. 3h**). In contrast, nearly all clonal overlap in UC was between Prlf. T_H_17.1 cells and T_H_17 cells, reinforcing the likelihood that actively proliferating T_H_ cells in UC are primarily of T_H_17 origin. We also noted *CD8A* expression in the CD4+ T_H_17 cell state (**Supplemental Fig. 3i**). Given our previous investigation demonstrating expansion of CD8+ T_H_17 cells in adult UC^8^, we sought to determine their activity in pediatric UC. We found that dual *CD4*+*CD8A+* T_H_17 cells were significantly enriched for clonal overlap with proliferating T_H_17.1 cells, suggesting active clonal expansion of this cell substate in UC (**Supplemental Fig. 3j**).

### T_H_1, T_H_17, and PD1Hi T_FH_ cells exhibit distinct cell state correlation networks

To explore the extent to which other immune pathways interacted with T_H_1, T_H_17, and PD1Hi T_FH_ populations, we performed a cell state abundance correlation analysis between these three CD4 subpopulations and each of the other annotated cell states (**Fig. 3a, Supplemental Table 9**). Our analysis revealed several cell states across all cell types that were significantly associated with each of these CD4 subpopulations. T_H_1 cells were associated with NK cells, inflammatory macrophages, and *TFPI2*-high fibroblasts, consistent with a type 1 immune response^37,38^. Amongst T_H_17 cells, significant correlations were observed for a variety of cell states, with the strongest correlations between MNPs and venules, as well as between T_H_17 cells and the *PI3*+ inflamed epithelium or *PDPN*+ remodeling fibroblasts. Importantly, while both T_H_17 and PD1Hi T_FH_ cells were enriched in UC patients, their correlation networks were largely distinct, with PD1Hi T_FH_ being primarily associated with IgG_1_ plasma cells, GC-like and proliferating B cells, and plasmablasts, consistent with their role in T cell-mediated B cell immune activation.

**Fig. 3.**
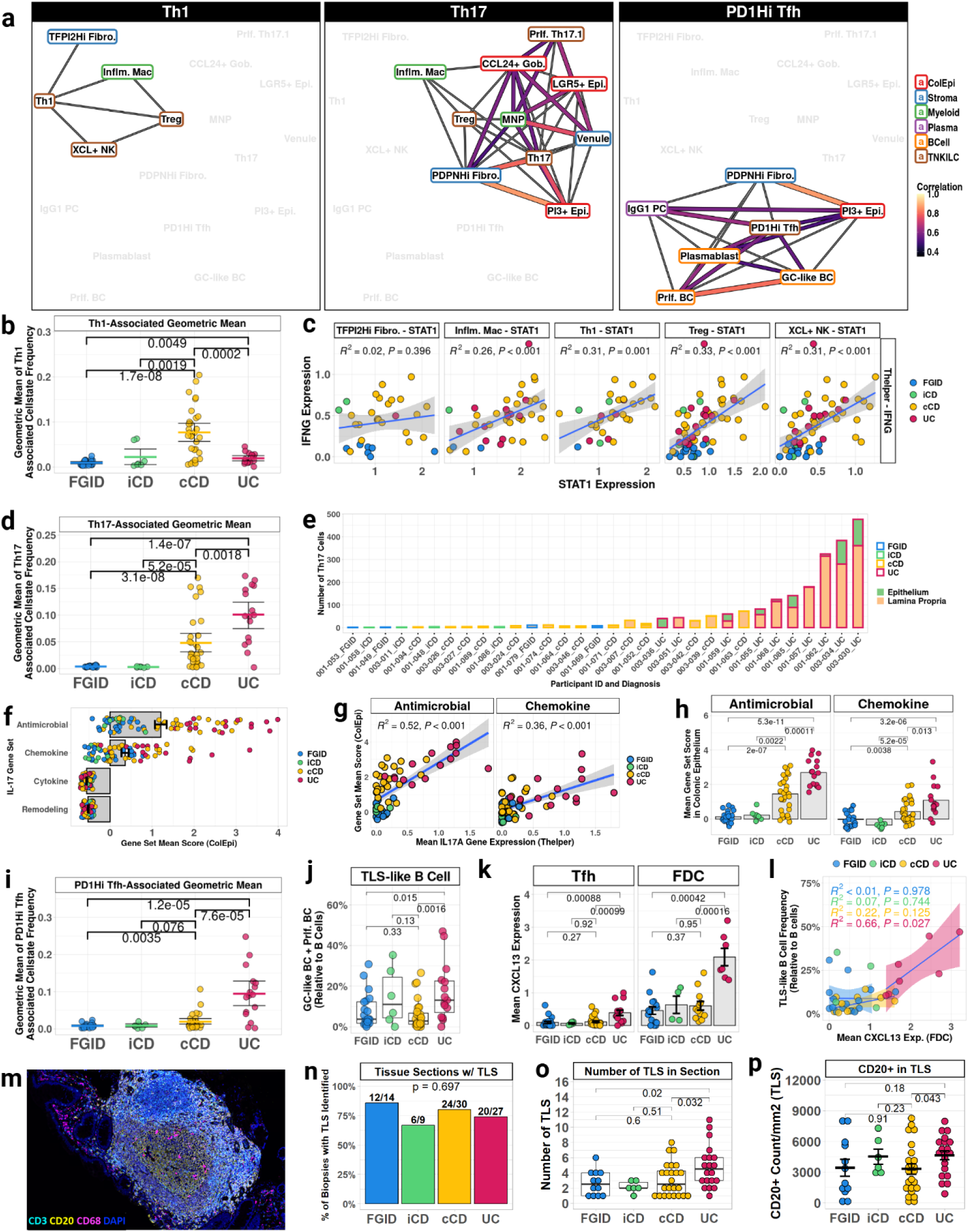
Crohn’s disease is characterized by T_H_1 and T_H_17-associated cellular networks; ulcerative colitis is characterized by T_H_17- and PD1Hi T_FH_-associated cellular networks. **a**, Cell state frequency Pearson correlation between T_H_1, T_H_17, and PD1Hi T_FH_ cell state and all other cell states with ≥ 100 cells (IgD and IgE plasma cells excluded). Cell state frequencies calculated as number of cells in the cell state relative to number of cells in cell type, per diagnostic biopsy (FGID, n = 23. iCD, n = 7. cCD, n = 31. UC, n = 16). Cell types labeled by color (colonic epithelium = red; stroma = blue; myeloid cells = green; plasma cells = purple; B cells = orange, T cells, NK cells, and innate lymphoid cells = brown). Pearson correlation and statistical testing performed on per-biopsy cell state frequencies. Cell state frequencies associated with T_H_1, T_H_17, and PD1Hi T_FH_ frequency at p < 1e-3 are visualized as force-directed network plot. Edges are shown if Pearson correlation ≥ 0.35. Edges gray if 0.35 < Pearson correlation < 0.5, edges colored if Pearson correlation ≥ 0.5. Width and color of edges correspond to strength of Pearson correlation ≥ 0.5. **b**, Geometric mean analysis of cell state frequencies associated with T_H_1 cell state, compared among disease categories. Cell state frequencies and biopsies used as input for correlation analysis also used as input for geometric mean analysis. **c**, Relationships between T_H_ cell subtype expression of *IFNG* and T_H_1-associated cell states, modeled as simple linear regressions. **d**, Geometric mean analysis of cell state frequencies associated with T_H_17 cell state, performed as in (**b**). **e**, Number of T_H_17 cells in EPI (green) and LP (beige) biopsy fractions, per biopsy with successfully processed LP and EPI fractions, ordered by total number of T_H_17 cells. Disease group is denoted by bar outline color. Study participant ID and disease group are labeled on x-axis. **f**, IL-17 gene set scores in diagnostic biopsies with at least 10 colonic epithelial cells (n = 77). **g**, Relationship between per-biopsy mean expression of *IL17A* in Thelper cells and IL-17 antimicrobial and chemokine gene set scores in colonic epithelial cells, modeled as a simple linear regression among diagnostic biopsies with at least 10 Thelper cells and 10 colonic epithelial cells (n = 66). **h**, Mean antimicrobial and chemokine gene set score in diagnostic biopsies with at least 10 colonic epithelial cells (FGID, n = 23. iCD, n = 7. cCD, n = 31. UC, n = 16), compared across disease groups. **i**, Geometric mean analysis of cell state frequencies associated with PD1Hi T_FH_ cell state, performed as in (**b**). **j**, Frequency of GC-like B cells and proliferating B cells combined into single group (“TLS-like B Cells”), relative to all B cells, per diagnostic biopsy with at least 10 B cells (FGID, n = 21. iCD, n = 6. cCD, n = 31. UC, n = 15). **k**, Mean T_FH_- and FDC-derived *CXCL13* log-normalized expression among diagnostic biopsies with ≥ 10 T_FH_ cells (FGID, n = 20; iCD, n = 5; cCD, n = 29; UC, n = 14) or FDC cells (FGID, n = 15; iCD, n = 4; cCD, n = 12; UC, n = 7). **l**, Relationship between FDC expression of *CXCL13* and frequency of TLS-like B cells, modeled as a simple linear regression within each diagnosis category. Biopsies included for analysis contained ≥ 10 FDCs and ≥ 10 B Cells (FGID, n = 15; iCD, n = 4; cCD, n = 12; UC, n = 7). iCD confidence interval not displayed for clarity. **m**, Representative image from colon of IBD participant with lymphoid aggregate, stained for CD3 (cyan), CD20 (yellow), CD68 (magenta), DAPI (dark blue). **n**, Percentage of tissue sections with any TLS identified. Chi-squared test used for statistical analysis. **o**, Number of TLS annotated per tissue section with any TLS identified, comparing FGID (n = 12, 7 participants), iCD (n = 6, 5 participants), cCD (n = 24, 15 participants), UC (n = 20, 11 participants). **p**, Density of CD20+ cells in TLS per tissue section with any TLS identified. All error bars denote standard error of the mean. Mann-Whitney U test, two-tailed, used for all pairwise comparisons. Shaded areas of regressions show 95% confidence interval. TLS, tertiary lymphoid structure.

### The T_H_1 correlation network is enriched in cCD and associated with STAT1-high cell states

We calculated the per-biopsy geometric mean of the cell states involved in each correlation network to determine the extent to which each network aligned with each disease state. We found that cCD was distinct from FGID, UC, and iCD in its enrichment for T_H_1-associated cell states (**Fig. 3b**). Because IFNγ is a predominant T_H_1 cytokine, we next determined whether IFNγ may be associated with the cell states in the T_H_1 network. We first examined *STAT1* expression in all cell states, given that *STAT1* both mediates and is induced by IFNγ signaling^39,40^. We found that 4 of the 6 top *STAT1*-expressing cell states (Inflm. Mac, T_H_1, MNP, TFPI2Hi Fibro.) were present in the T_H_1 correlation network (**Supplemental Fig. 4a**). We then compared *IFNG* expression of *IFNG*-expressing cell subtypes (including T_H_, NK, CTL, and InvariantT; **Supplemental Fig. 4b-c**) to *STAT1* expression in T_H_1-associated cell states within the same biopsy, finding significant correlations between T_H_ *IFNG* expression and *STAT1* expression in inflammatory macrophages, T_H_1 cells, T_regs_, and *XCL*+ NK cells (**Fig. 3c**, **Supplemental Table 10**). The specificity of this correlation was underscored by the fact that *IFNG* from other *IFNG*-expressing cell subtypes was not associated with *STAT1* in any T_H_1-associated cell states. (**Supplemental Fig. 4d**). These correlations were reproduced when we restricted the dataset from all enrolled patients to focus only on cCD and UC biopsies, suggesting that the T_H_ IFNγ association with *STAT1* expression is not solely due to the overall status of tissue inflammation (**Supplemental Fig. 4e**).

### The T_H_17 correlation network is most substantially enriched in UC, and is associated with epithelial antimicrobial and chemokine response

Although cCD was significantly enriched for T_H_17-associated states compared to both FGID and iCD, this network was most enriched in UC patients (**Fig. 3d**). To further interrogate T_H_17 signaling, we next focused on IL-17A, the key T_H_17 cytokine. IL-17A is known to have multiple effects, including enhanced recruitment of peripheral leukocytes and increased antimicrobial defense^36,41–43^. We therefore created gene set scores for IL-17-induced antimicrobial function, chemokine production, cytokine production, and tissue remodeling using genes downstream of IL-17 signaling in the KEGG IL-17 signaling pathway (**Supplemental Fig. 5a**)^44–46^. These gene sets were active across multiple cell types (**Supplemental Fig. 5b**, **Supplemental Table 11**). Given IL-17 effects on barrier function and the presence of T_H_17 cells within the epithelial fraction of our biopsies (**Fig. 3e**, **Supplemental Fig. 1a**), we focused our analyses on IL-17 effects in colonic epithelium. The antimicrobial and chemokine gene sets were active in colonic epithelium (**Fig. 3f**). We found strong associations between *IL17A* in T_H_ cells and activity of IL-17 gene sets in colonic epithelium, suggesting either a direct influence of *IL17A* on this epithelium, or a common process contributing to expression of these genes in distinct cell types (**Fig. 3g**). Strong correlations remained when we restricted analysis to cCD and UC biopsies only, but the correlations were not observed between *IL17A* and active IL-17 gene sets in other cell types (**Supplemental Fig. 5c**). The antimicrobial and chemokine gene sets, and associated antimicrobial and chemokine genes, were active in similar colonic epithelial cells, were significantly enriched in cCD compared to FGID, and were further enriched in UC (**Fig. 3h**, **Supplemental Fig. 5d-g**). Because many of the chemokines in the IL-17-induced chemokine gene set are known neutrophil chemoattractants, we contextualized the scRNA-seq results with multiplexed IHC and found profound neutrophilia in UC epithelium (**Supplemental Fig. 5i**). A similar increase in potentially inflammatory conventional CD4 T cells (CD3+CD8-FOXP3-), was not observed histologically in UC epithelium relative to cCD (**Supplemental Fig. 5j**).

### The PD1Hi T_FH_ correlation network is enriched in UC and is associated with generation of humoral immunity

In addition to their enrichment in T_H_17 networks, UC patients also demonstrated significant enrichment in the PD1Hi T_FH_ network (**Fig. 3i**). This group of differentially abundant cell states was highly suggestive of increased T cell-dependent B cell activation and tertiary lymphoid structure activity. The frequencies of GC-like B cells and proliferating B cells were highly correlated (R = 0.8) (**Fig. 3a, Supplemental Table 9**). We considered that these cell states may be part of tertiary lymphoid structures (TLS). We thus combined both cell states into a “TLS-like B Cell” annotation and observed this grouping of cells to be increased in UC biopsies relative to both FGID and cCD (**Fig. 3j**). We then examined expression of the germinal center-organizing cytokine *CXCL13*. Its expression was increased in UC T_FH_ cells and was greatly enriched in UC follicular dendritic cells (FDCs) (**Fig. 3k**). Notably, *CXCL13* expression among FDCs correlated with TLS-like B cell frequency, but only in UC (**Fig 3l**). Histologically, we identified lymphoid aggregates of B and T cells, which contained a center of CD20+ cells surrounded by a ring of CD3+ cells (morphologically resembling a germinal center), in biopsies across all disease groups (**Fig. 3m-n**). Importantly, amongst biopsies with any identified TLS, we observed an increase in the number of TLS per biopsy in UC, as well as increased CD20+ cell density within these structures in UC relative to cCD (**Fig. 3o-p**).

### Predictors of Anti-TNF Response in IBD: cCD

#### Differential gene expression at diagnosis does not distinguish anti-TNF treatment or response groups in cCD

Because anti-TNF agents are the first line (and only FDA-approved) biologic therapy in pediatric IBD, we next investigated whether we could determine differential expression signatures associated with response to these agents from data in diagnostic biopsies (obtained prior to any treatment) from IBD patients. We first performed per-cell type differential expression (DE) analysis between cCD patients who would be treated with anti-TNF and those not treated with anti-TNF in the 2-year follow-up period (the ‘NOA’ patients). This analysis identified a paucity of DE differences, with the only DE gene identified being *TNFAIP6*, for which the enrichment was largely driven by a single outlier sample (**Supplemental Fig. 6a-b, Supplemental Table 6**). We next performed a similar DE analysis to compare cCD FR to PR patients at the time of diagnostic biopsy. This analysis identified only the plasma cell gene *PHGDH* (**Supplemental Fig. 6c-d, Supplemental Table 6**) differentially expressed between the two groups. This enzyme, critical for serine synthesis, was enriched in cCD PR vs. FR^47^. These results underscore the challenge in creating a predictive signature at diagnosis for anti-TNF response in cCD^48,49^.

#### Post-anti-TNF treatment cCD biopsies are defined by a decrease in IFNG signaling

Unlike the lack of differences in diagnostic cCD biopsies, an immunologic signature did emerge when post-treatment cCD biopsies were compared between FR and PR patients. To determine the cell types most likely to reflect an immunologic impact from anti-TNF therapies, we first scored each cell type for pre-treatment TNF pathway activity (**Fig. 4a**). Both myeloid cells and TNKILCs demonstrated elevated TNF pathway scores and were thus chosen for more detailed analyses. We found that TNF pathway scores were decreased for both myeloid cells and TNKILCs in post-treatment cCD relative to pre-treatment for both response groups, consistent with a biologic impact of anti-TNF on all treated patients (regardless of anti-TNF response) (**Fig. 4b-c**)^50,51^. To more deeply interrogate the molecular impact of anti-TNF on myeloid and TNKILCs, we conducted a DE analysis in these cell types, comparing pre- and post-treatment cCD, without regard to treatment response (**Fig. 4d**). Several immune genes were found to be upregulated in pre-treatment cCD myeloid cells and TNKILCs relative to post-treatment, including *STAT1*, cytolytic genes (*GNLY*, *PRF1*), calprotectin (*S100A8*, *S100A9*), and IFNγ-induced genes (*CXCL9*, *CXCL10*, *GBP1*, *GBP2*, *GBP5*)^52^. Moreover, “Interferon gamma signaling” was the top Reactome term enriched in pre-treatment cCD differential expression, consistent with IFNγ-induced inflammatory states suggested in the T_H_1 network (**Fig. 3c**, **4e**, **Supplemental Table 6**). Based on these findings, we compared pre- and post-treatment cell state frequencies of T_H_1-associated cell states among cCD biopsies, finding decreases in each cell state (**Fig. 3a, 4f**). Additionally, the two other top *STAT1*-expressing cell states, MNPs and T_H_17 cells, were also decreased in post-treatment cCD, when all treatment response groups were analyzed together (**Fig. 4f**). Overall, these findings suggest successful abrogation of IFNγ signaling through anti-TNF therapy in cCD, regardless of clinical treatment response.

**Fig. 4.**
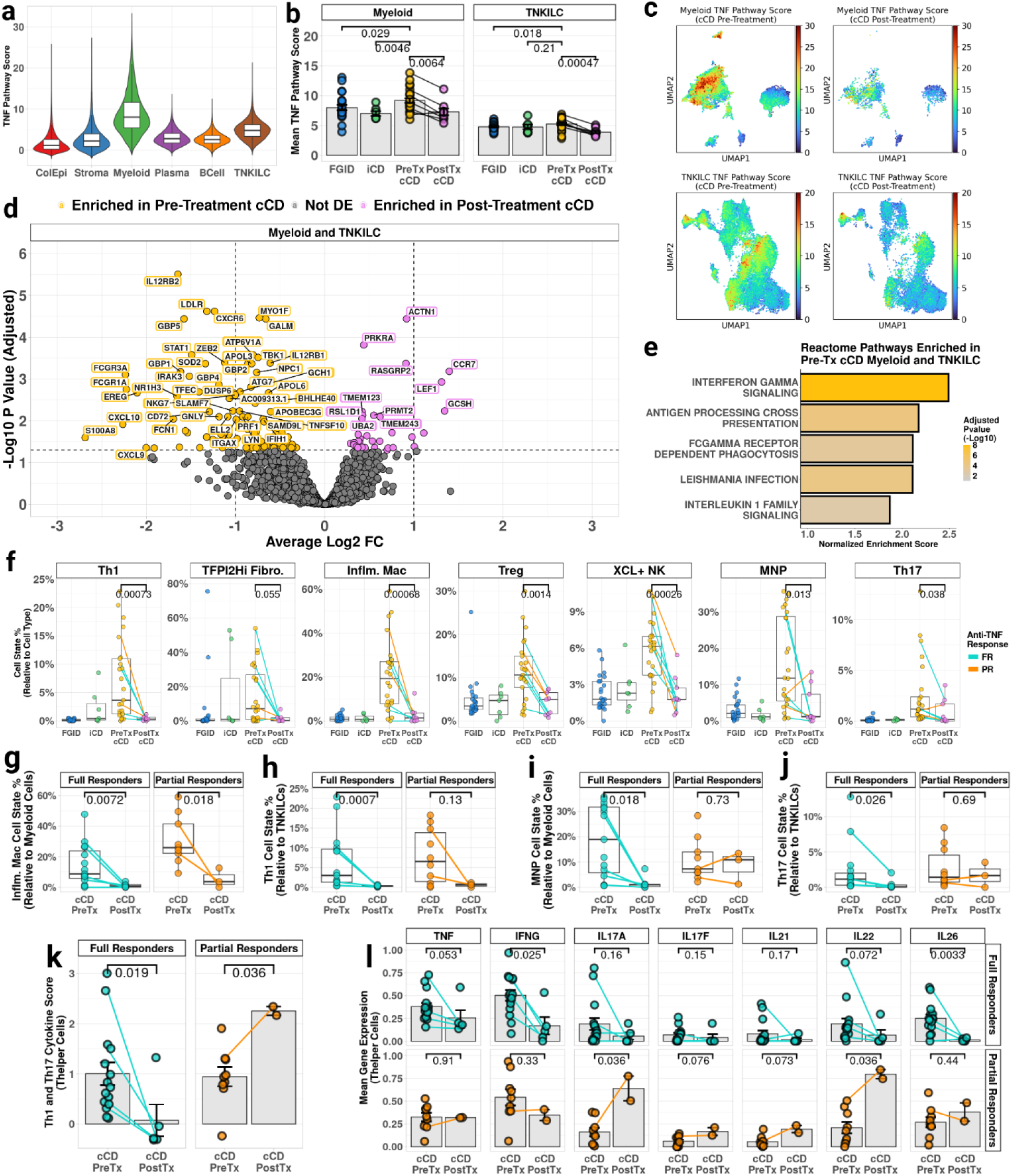
Anti-TNF decreases T_H_1-associated cell states, but T_H_17-associated cytokine expression is persistent in post-treatment cCD partial responders a, Violin plot of TNF pathway scores in all cell groups. Cells were scored using decoupler multivariate linear model. PROGENy “TNFa” pathway genes and weights were used as input, filtered to genes with p-value < 1e-10 (225 genes). **b,** Pre- and post-treatment TNF scores in cCD, excluding cCD participants who did not receive anti-TNF. FGID and pre-treatment iCD included as reference. Biopsies used had ≥ 10 cells in each cell type (Myeloid: FGID, n = 22; iCD, n = 7; Pre-Tx cCD, n = 24; Post-Tx cCD, n = 9. TNKILC: FGID, n = 23; iCD, n = 7; Pre-Tx cCD, n = 25; Post-Tx cCD, n = 9). **c**, UMAP of TNF pathway score for myeloid (top) and TNKILC (bottom) cell groups among pre-treatment (left) and post-treatment (right) cCD biopsies. **d,** Differential expression analysis among myeloid cells and TNKILCs, comparing pre-treatment and post-treatment cCD (Pre-tx, n = 25; Post-tx, n = 9). Pseudobulk differential expression calculated with DESeq2. Genes labeled if adjusted p-value ≤ 0.01. **e**, Enriched terms from gene set enrichment analysis of differential expression between pre- and post-treatment cCD patients. Gene ranks computed using DESeq2 Wald test “stat” value (log2FC/SE(log2FC)). Ranked genes were tested using Reactome 2023 v2 gene sets with 15 ≤ number of genes ≤ 100 and sorted for normalized enrichment score. Terms enriched in pre-treatment cCD with the 5 lowest adjusted p-values are shown. **f**, Changes in cell frequencies among T_H_1-associated cell states, as well as MNP and T_H_17 cell states (FGID, n = 23; iCD, n = 7; Pre-Tx cCD, n = 24 for stromal and myeloid cell states, n = 25 for TNKILC cell states; Post-cCD, n = 9). **g-j**, pre- and post-treatment cell frequency of inflammatory macrophages (**g**), T_H_1 cells (**h**), mononuclear phagocytes (**i**), and T_H_17 cells (**j**), relative to their respective cell type, among cCD full responder and partial responder biopsies with at least 10 cells in relevant cell type (cCD FR Pre, n = 15; cCD FR Post, n = 6; cCD PR Pre, n = 9 for Myeloid, n = 10 for TNKILC; cCD PR Post, n = 3). **k**, Mean module score of canonical T_H_1 and T_H_17 cytokine gene expression (module input genes: *IFNG*, *TNF*, *IL17A*, *IL17F*, *IL21*, *IL22*, *IL26*), grouped into timepoint and response categories for cCD biopsies with at least 10 T_H_ cells (cCD FR Pre-Tx, n = 15; cCD FR Post-Tx, n = 5; cCD PR Pre-Tx, n = 9; cCD PR Post-Tx, n = 2). **l**, Mean log-normalized expression of genes used as input for module score in (**k**) among T_H_ cell subtype, grouped into timepoint and response categories for cCD biopsies with at least 10 T_H_ cells. All error bars denote standard error of the mean. Mann-Whitney U test, two-tailed, used for pairwise comparisons. Lines connect pre- and post-treatment biopsies from the same study participant.

#### Post-treatment cCD partial responders retain T_H_17 signaling

With overall differences in pre- and post-treatment cCD delineated, we investigated whether post-treatment biopsies would illuminate differences in cCD FR and cCD PR. This analysis revealed decreases in the T_H_1-associated inflammatory macrophage cell state in both FR and PR (**Fig. 4g**), and that T_H_1 cells themselves were significantly decreased in FR with a trend towards reduction in PR (**Fig. 4h**). In contrast, the T_H_17-associated MNP cell state (shown in **Fig.3a**), as well as T_H_17 cells themselves, were decreased only in cCD FR, while being maintained in PR (**Fig. 4i-j**). To more fully evaluate canonical T_H_1 and T_H_17 cytokine gene expression, we analyzed the expression of key T_H_1 cytokine genes (*IFNG*, *TNF)* and T_H_17 cytokine genes (*IL17A, IL17F*, *IL21*, *IL22*, *IL26)* within the T_H_ cell subtype. Consistent with the findings above, a gene signature created from these cytokines was *significantly reduced* in the T_H_ cells from cCD FR patients post-treatment relative to pre-treatment, but *significantly increased* in cCD PR patients post-treatment T_H_ cells relative to pre-treatment (**Fig. 4k**)^50^. Single gene analysis of pre- and post-treatment T_H_ cells revealed trends toward reduction in *TNF*, *IFNG*, *IL22*, and *IL26* in cCD FR (**Fig. 4l**). In contrast, cCD PR T_H_ cells demonstrated unchanged *TNF* and *IFNG*, along with trends towards increases in *IL17A*, *IL17F*, *IL21*, and *IL22* after anti-TNF treatment (**Fig. 4l**). Together, these data point to persistent T_H_17 immunopathology associated with anti-TNF PR in cCD.

### Predictors of Anti-TNF Response in IBD: UC

#### UC diagnostic biopsies from anti-TNF-treated patients are enriched for markers of inflammation and epithelial cell cycling

In the PREDICT cohort, the number of UC patients with repeat biopsies (2/16) was too few to rigorously define the post-treatment impact of anti-TNF therapies on immune cells and interactions. However, unlike for CD, UC diagnostic biopsies were discriminative of anti-TNF response. To discover this, we first performed an analysis comparing diagnostic biopsies from UC patients later treated with anti-TNF to NOA patients, utilizing the per-cell type DE analysis paradigm described above. Unlike in cCD patients, where informative gene expression changes were not observed, in UC, several notable genes were transcriptomically distinct in patients for whom the treating physician initiated anti-TNF therapy versus the NOA patients. Among colonic epithelial cells in pre-treatment biopsies, genes enriched in UC NOA patients were notable for *CA1*, *CA2*, and *SLC26A3*, representing normal colonic absorptive function (**Supplemental Fig. 7a**, **Supplemental Table 12**). Genes enriched in the colonic epithelium from UC patients who would eventually be treated with anti-TNF therapies included cell cycle-associated genes, such as *RRM2*, *ZWINT*, and *CENPM* (**Supplemental Fig. 7a**, **Supplemental Table 12**). Reactome pathways enriched for the DE results included GTPase cell signaling pathways for UC NOA patients, and DNA synthesis and replication pathways for UC anti-TNF patients (**Supplemental Fig. 7b-c**). Additional notable DE genes in UC patients treated with anti-TNF therapies included the neutrophil chemoattractant *CXCL6* in stromal cells, and the inflammatory cytokine-induced ferritin transport genes *FTL* and *FTH1* in myeloid cells^53,54^ (**Supplemental Fig. 7a**, **Supplemental Table 6**).

#### At the time of diagnostic biopsy, UC patients who would become anti-TNF NR were enriched for extracellular remodeling and T cell differentiation genes

We next evaluated diagnostic biopsies from UC patients who were eventually treated with anti-TNF for per-cell type DE that could differentiate PR versus NR. Of note, PR was the best response achieved in the PREDICT UC cohort (although other studies have documented UC patients achieving FR to anti-TNF)^55–57^. In patients that partially responded to anti-TNF versus NR patients, pre-treatment UC colonic epithelium and TNKILCs were enriched for genes in cellular metabolism and maintenance pathways (**Fig. 5a-b**). *MIF*, which promotes *TNF* induction and whose release is induced by microbial proteins, was enriched in UC PR versus NR myeloid cells, B cells, and TNKILCs (**Fig. 5a**)^58–60^. Additional notable genes enriched in UC PR versus NR included *LYZ* (an antimicrobial enzyme) in myeloid cells and interferon-stimulated genes *IFI6*, *IFITM1*, and *ISG15* in plasma cells (**Fig. 5a**)^61^.

**Fig. 5.**
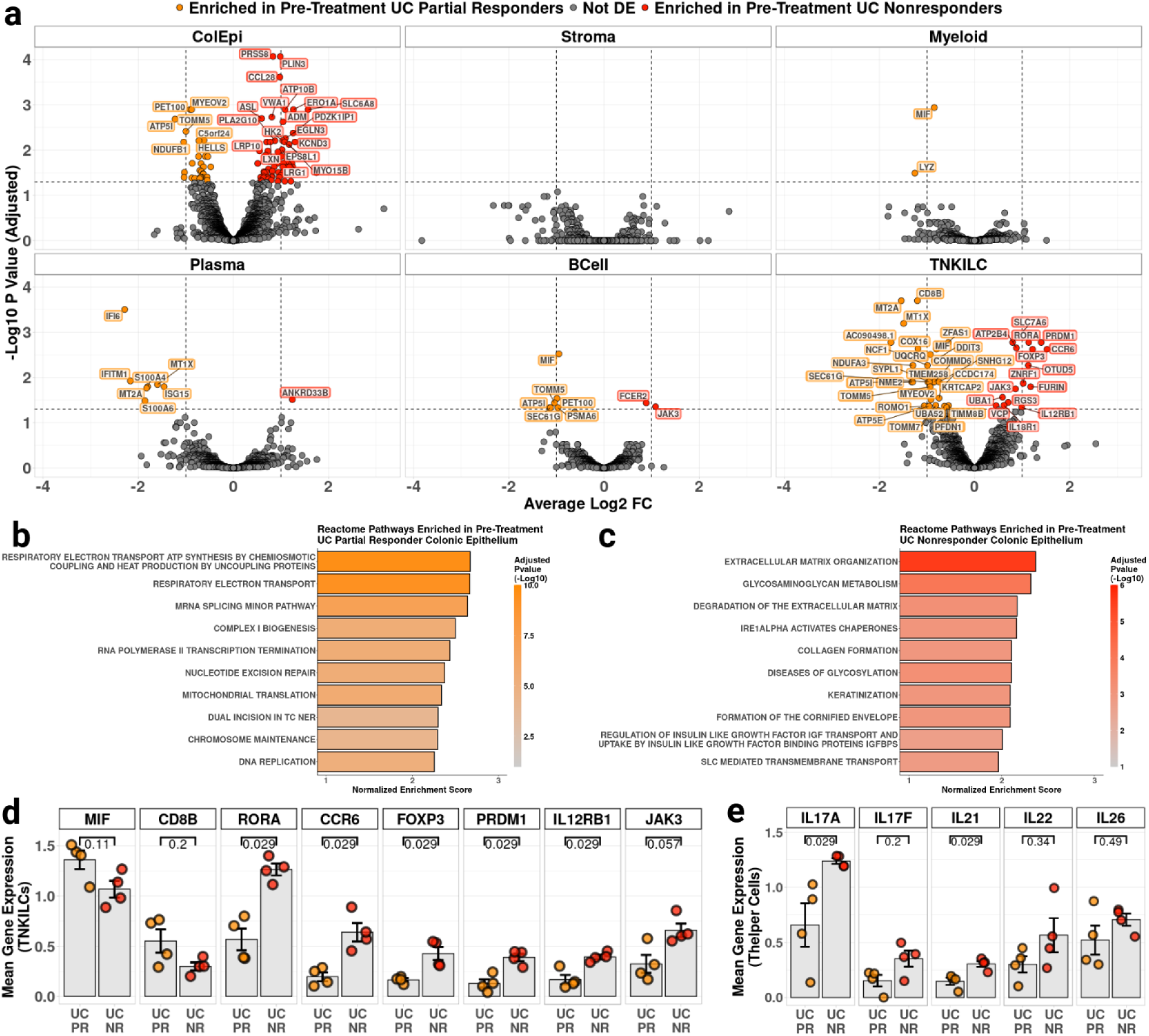
*IL17A* expression is enriched in UC anti-TNF nonresponders prior to treatment **a**, Differential expression comparing differentially expressed genes in post-treatment UC full responders and partial responders within each annotated cell type. 4 PR and 4 NR post-treatment biopsies compared. Genes used as input determined through filtering by expression in a minimum number of samples. Genes shown if adjusted p value ≤ 0.05, except for ColEpi (ColEpi DE genes shown if adjusted p value ≤ 0.01). **b**, **c**, Enriched terms from gene set enrichment analysis of differential expression between pre-treatment colonic epithelium from UC PR (**b**) and NR (**c**) participants. Top 10 terms for each treatment group shown. Gene ranks computed using DESeq2 Wald test “stat” value (log2FC/SE(log2FC)). Ranked genes tested using Reactome 2023 v2 gene sets with 15 ≤ number of genes ≤ 100 and sorted for normalized enrichment score. **d**, Mean log-normalized expression of selected DE genes among TNKILCs, grouped into response categories for pre-treatment UC biopsies with at least 10 TNKILC cells (UC PR, n = 4; UC NR, n = 4). **e**, Mean log-normalized expression of selected T_H_17-associated cytokines among T_H_ cell subtype, grouped into response categories for pre-treatment UC biopsies with at least 10 T_H_ cells (UC PR, n = 4; UC NR, n = 4). All error bars denote standard error of the mean. Mann-Whitney U test, two-tailed, used for pairwise comparisons.

In contrast, UC NR versus PR colonic epithelium demonstrated enrichment in pathways governing extracellular degradation and remodeling (**Fig. 5a, c**). Notably, *FCER2* (encoding CD23) and *JAK3* (induced upon CD40 ligation or bacterial stimulation, necessary for lymphocyte proliferation and differentiation, and for which there is an FDA-approved therapeutic in adult UC) were enriched in UC NR B cells^62–65^. Many of the genes enriched in UC NR TNKILC are involved in differentiation, activation, and migration of CD4 T cells, including *RORA* (T_H_17 cytokine production), *CCR6* (T_H_17 and T_reg_ migration), *FOXP3* (T_reg_ differentiation), *PRDM1* (encoding BLIMP-1, which mediates chronic activation/exhaustion), *IL12RB1* (subunit of IL-12 and IL-23 receptors), and *JAK3* (**Fig. 5a**)^66–71^. We also examined per-biopsy expression of several TNKILC DE genes in the TNKILC cell type, which recapitulated the DE findings described above (**Fig. 5d**). Of note, in addition to therapeutics directed at *JAK3*, *IL12RB1* is involved in pathways targeted by anti-IL-12p40/anti-IL23p19 therapies, underscoring the potential availability of alternatives to anti-TNF in UC patients bearing anti-TNF nonresponse markers^64,72^.

#### At the time of diagnostic biopsy, UC patients who would become anti-TNF NR were enriched for T_H_ IL17A

We further evaluated the role of T_H_17 cytokine signaling in defining UC treatment response. We found that, compared to pre-treatment UC PR, UC NR T_H_ cells were enriched in T_H_17 cytokines, with all UC NR biopsies having greater T_H_ *IL17A* and *IL21* than UC PR biopsies (**Fig. 5e**). Thus, *IL17A* was identified as a marker of anti-TNF therapy failure in both cCD and UC.

#### A distinct T peripheral helper (T_PH_) subset was not detected in pediatric IBD patients

In addition to T_H_17 cells, adult UC has previously been associated with T_PH_ cells^26^. Thus, we explored whether T_PH_ cells were present in the pediatric IBD patients profiled in PREDICT. We found that neither the PD1Lo T_FH_ nor the PD1Hi T_FH_ population clearly expressed T_PH_ markers (**Supplemental Fig. 8a-b**, **Supplemental Table 7**)^73^. Additionally, no clear subpopulation of cells expressed a combination of canonical T_PH_ markers^73^, assessed through UMAP visualization and through gene expression across a pseudotime diffusion vector spanning CD4 T cells (**Supplemental Fig. 8c-e**, **Supplemental Table 7**). This may represent a phenotypic difference in adult and pediatric UC.

#### UC T_H_17 cells exhibit T_FH_ features and are inversely correlated with a UC-specific shift towards IgG_1_ plasma cells

While no evidence for an association between T_PH_ and UC patients was observed, we did identify a subset of T_FH_ cells that were altered in PREDICT’s UC patients: We found that although total T_FH_ cells were present in similar relative frequencies in all PREDICT disease groups, UC could be distinguished by the fact that T_FH_ cells in these patients were more shifted towards the PD1Hi T_FH_ cell state (**Fig. 6a-b**). Moreover, amongst T_FH_ genes involved in B cell help^73^, we found *CXCL13* to be increased in UC PD1Hi T_FH_ relative to cCD (**Fig. 6c**, **Supplemental Fig. 8a**). During cell state annotation, we also observed that B cell help genes were present in T_H_17 cells (**Supplemental Fig. 2f**), with *MAF*, *PDCD1*, *IL21*, and *CXCL13* being increased in UC T_H_17 cells compared to cCD T_H_17 cells (**Fig. 6d**).

**Fig. 6.**
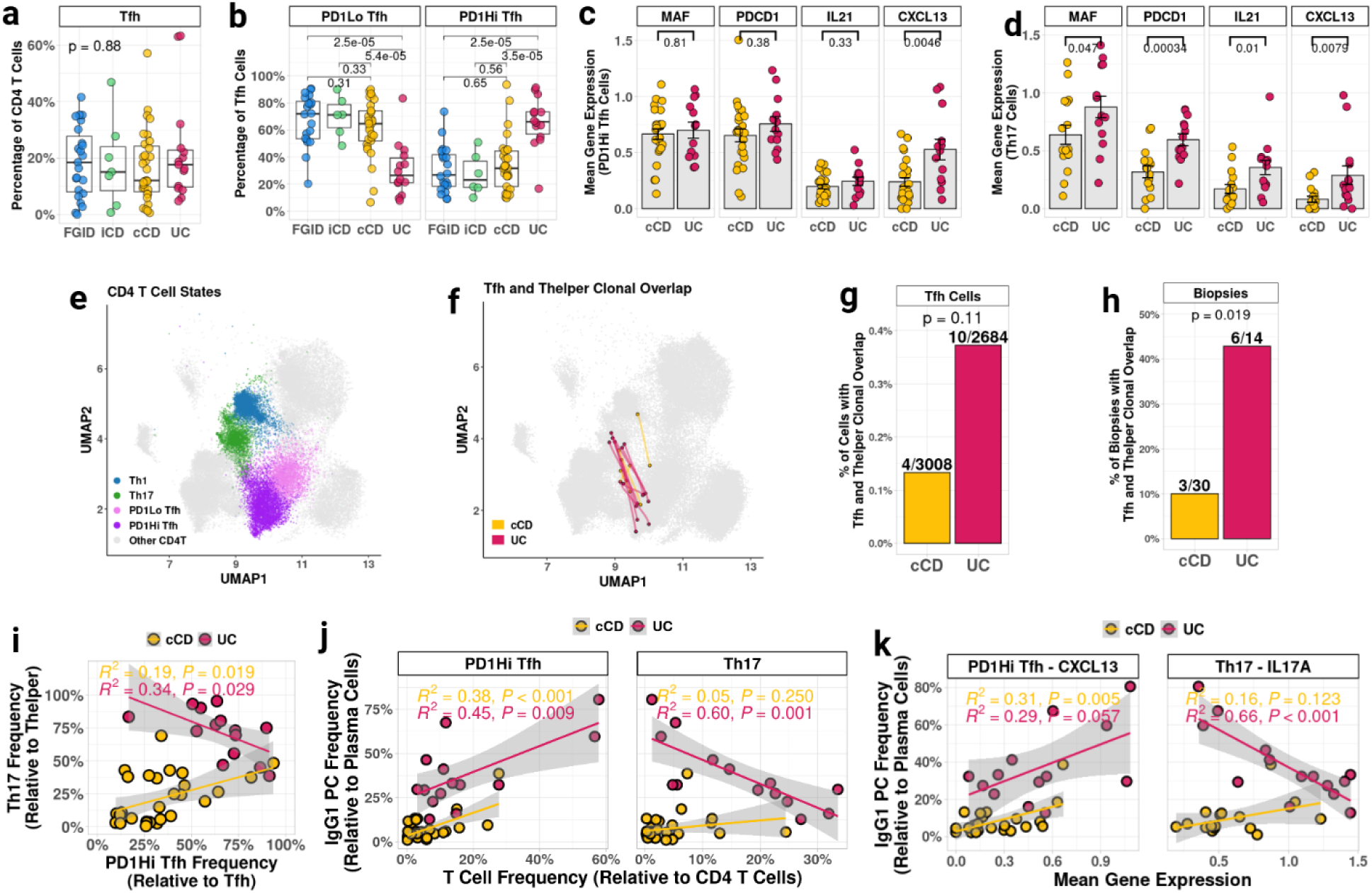
T_H_17 and PD1Hi T_FH_ cells belong to shared clonotypes yet are inversely correlated in UC. **a**, Frequencies of T_FH_ cells relative to all CD4 T cells, per diagnostic biopsy with ≥ 10 CD4 T cells (FGID, n = 23; iCD, n = 7; cCD, n = 31; UC, n = 15). Kruskal-Wallis test used for statistical testing. **b**, Frequencies of T_FH_ cell states relative to all T_FH_ cells, per diagnostic biopsy with ≥ 10 T_FH_ cells (FGID, n = 20; iCD, n = 6; cCD, n = 29; UC, n = 14). **c**, **d**, Mean log-normalized per-biopsy gene expression of canonical T_FH_ B cell help genes in PD1Hi T_FH_ cells (**c**) or T_H_17 cells (**d**), per cCD and UC diagnostic biopsy with ≥ 10 cells in cell state (PD1Hi T_FH_: cCD, n = 25; UC, n = 13. T_H_17: cCD, n = 16; UC, n = 14)). **e**, UMAP of CD4 T cell states. T_H_1, T_H_17, PD1Lo T_FH_, and PD1Hi T_FH_ cells highlighted. **f**, UMAP of shared clones between T_H_ and T_FH_ cells among T_H_1, T_H_17, PD1Lo T_FH_, and PD1Hi T_FH_ cell states in cCD (yellow lines), or UC (red lines) diagnostic biopsies. Shared clones determined by paired CDR3αβ amino acid sequence. **g**, Percentage of cCD and UC cells in diagnostic biopsies with any identified clonal overlap between T_H_ and T_FH_ cells, as a fraction of total T_FH_ cells within disease group. Fisher’s exact test was used to compare number of shared clones in cCD or UC cells. **h**, Percentage of cCD and UC diagnostic biopsies with any identified clonal overlap between T_H_ and T_FH_ cells, among biopsies with detected CDR3αβ in T_H_1, T_H_17, PD1Lo T_FH_, and PD1Hi T_FH_ cell states. Fisher’s exact test was used to compare presence or absence of any shared clones in cCD or UC biopsies. **i**, Linear regression between T_H_17 frequency and PD1Hi T_FH_ frequency, relative to their respective cell subtypes (T_H_ and T_FH_), within each disease group. Restricted to diagnostic cCD and UC biopsies with at least 10 cells in T_H_ and T_FH_ cell subtypes (cCD, n = 28; UC, n = 14). **j**, Frequency of IgG1 plasma cell relative to all plasma cells, compared to frequency of PD1Hi T_FH_ (left) or T_H_17 (right), relative to CD4 T cells, per cCD and UC diagnostic biopsies with ≥ 10 plasma cells and ≥ 10 CD4 T cells (cCD, n = 31; UC, n = 14). **k**, Linear regression between IgG_1_ plasma cell frequency and expression of *CXCL13* in PD1Hi T_FH_ cells and *IL17A* in T_H_17 cells. Restricted to diagnostic cCD or UC biopsies with at least 10 cells in the compared cell states (IgG1 PC vs. PD1Hi T_FH_: cCD, n = 24; UC, n = 13. IgG1 PC vs.T_H_17: cCD, n = 16; UC, n = 14). All unspecified per-biopsy grouped comparisons were statistically tested with Mann-Whitney U, two-tailed. All error bars denote standard error of the mean. Shaded areas of regressions show 95% confidence interval.

Given these data, we considered a model wherein UC T_H_17 cells, but not cCD T_H_17 cells, exhibit T_FH_-like plasticity. To test this model, we assessed which set of paired CDR3αβ sequences were detectable between non-proliferating T_FH_ cells and other T_H_ cells within a biopsy (**Fig. 6e-f**). Our analysis demonstrated that, in 6/14 UC diagnostic biopsies, there was evidence for T_FH_-T_H_ clonal overlap, while only 3/30 cCD diagnostic biopsies demonstrated T_FH_-T_H_ clonal overlap (**Fig. 6g-h, Supplemental Table 8**). Clonal sharing was primarily between T_H_17 and PD1Hi T_FH_, with only one clonal pair involving T_H_1 or PD1Lo T_FH_ cells (**Supplemental Table 8**). These data indicate that, although a small absolute number of shared clonotypes was identified, UC biopsies were enriched for identifiable T_FH_-T_H_ clonal overlap (**Fig. 6g-h**).

To determine whether the coordination of these phenotypic shifts would be distinct in cCD versus UC, we compared the relative frequencies of T_H_17 cells among all T_H_ cells, and PD1Hi T_FH_ cells among all T_FH_ cells. Notably, we found opposite patterns in the two IBD endotypes, with T_H_17 and PD1Hi T_FH_ cells being positively correlated in cCD, but inversely correlated in UC (**Fig. 6i**, **Supplemental Table 10**). Considering the implications of differences in T cell phenotypes on B cell help and the development of antibody-secreting cells, we explored whether PD1Hi T_FH_ cells or T_H_17 cells were associated with the shift towards IgG_1_ plasma cells observed in UC (**Fig. 2i**). We found that in both cCD and UC, PD1Hi T_FH_ cell proportion and *CXCL13* expression were positively correlated with IgG_1_ plasma cells (**Fig. 6j-k**). However, in T_H_17 cells, we found a *strong inverse correlation* of T_H_17 cell state frequency or *IL17A* expression and IgG_1_ plasma cells in UC, despite both T_H_17 and IgG_1_ plasma cells both being increased in UC compared to CD (**Fig. 2i, 2n, 6j-k**). These data suggest an antagonism between T_H_17-associated and T_FH_-associated signaling in UC.

#### UC T_FH_ TCRs exhibit polyreactivity signatures that are directly correlated with UC-specific plasma cell shifts and inversely associated with T_H_17-derived *IL17A*

Because of the prevalence of T-B interactions observed in UC, and known clinical associations with autoreactivity among IBD patients^74^, we hypothesized that UC TCRs may bear features that distinguish them from cCD TCRs, and that TCR phenotypes could play a role in the plasticity observed among UC T cells.

Previous studies have suggested that increased CDR3β hydrophobicity may predispose TCRs to polyreactivity or autoreactivity through non-specific interactions^75–77^. Pointing towards poly- and/or autoreactivity in UC T cells, we observed a relative increase in usage of large, aliphatic, and hydrophobic amino acid residues, with a corresponding decrease in polar amino acids, in the polymorphic middle region of CD4 T cell CDR3β sequences in patients with UC (**Fig. 7a-b**, **Supplemental Table 8**). To relate these changes to functional differences in TCRs, we applied the previously-defined middle-TCR intrinsic regulatory potential (mTiRP) score^78^. mTiRP is a TCRβ-based metric created from the association of CD4 TCRs with the T_reg_ phenotype, given that T_regs_ derived from the thymus are trained to recognize self and commensal antigens, and are thus more likely to be self-reactive^78,79^.

**Fig. 7.**
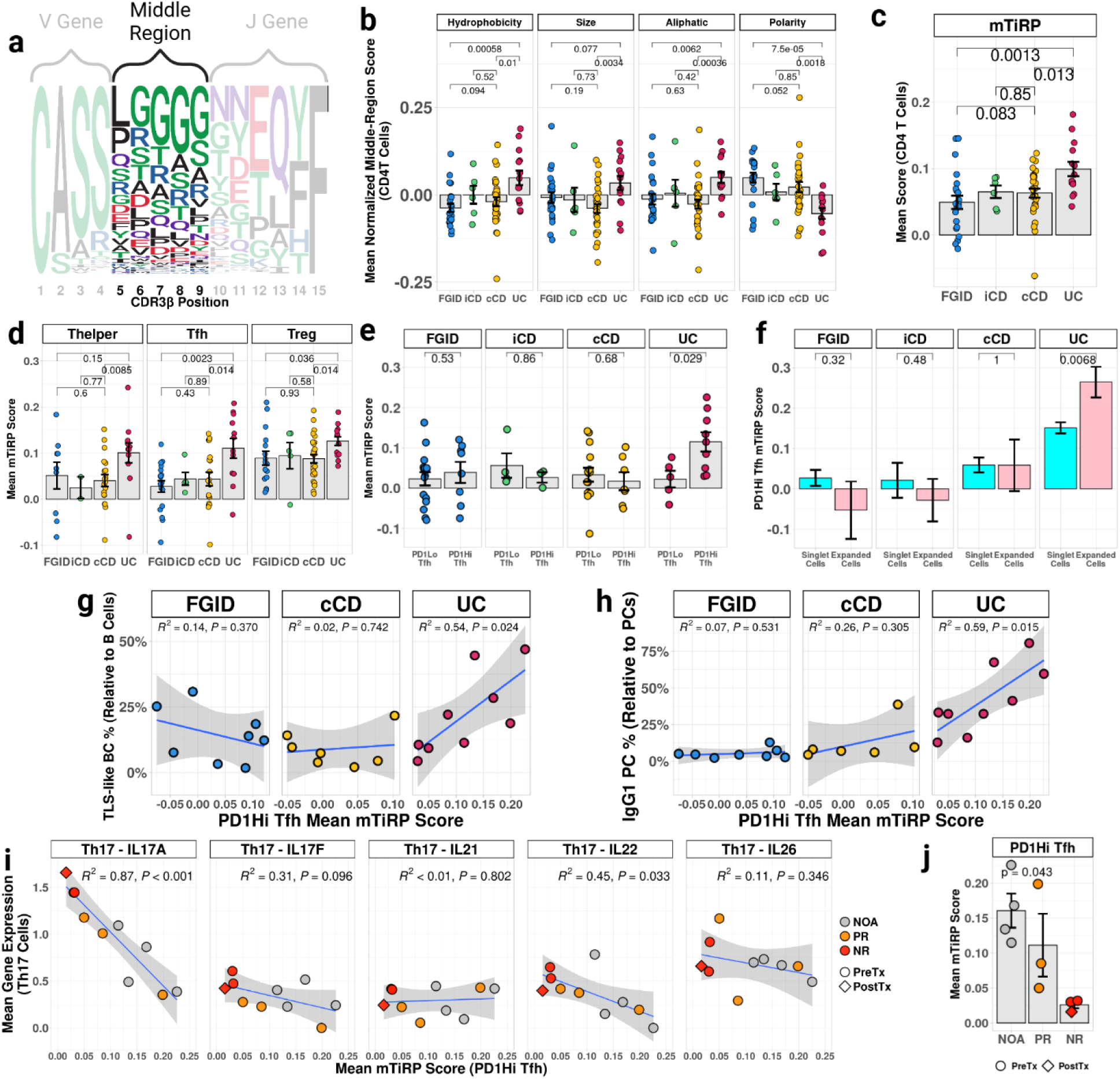
UC PD1Hi T_FH_ cells are enriched for polyreactive TCR features that are inversely correlated with *IL17A* gene expression. **a**, Logo plot of CDR3β amino acid usage among baseline CD4 T cells, for length 15 CDR3β sequences. Brackets denote areas of V gene contribution, J gene contribution, and N or P nucleotide contribution (middle region). **b**, CDR3β middle-region metrics of biophysical properties among diagnostic biopsies with ≥ 50 CD4 T cells with CDR3β detected (FGID, n = 22; iCD, n = 6; cCD = 30; UC = 14). Quantification of CDR3β middle-region biophysical properties calculated using R package alakazam and Gaussian-normalized within PREDICT dataset. Mean score denoted by gray bar. **c**, Mean mTiRP score in CD4 T cells among diagnostic biopsies with ≥ 50 CD4 T cells with CDR3β detected. **d**, Mean mTiRP score calculated among CD4 T cell subtypes T_H_, T_FH_, and T_reg_ within FGID, cCD, and UC diagnostic biopsies. Biopsies included have ≥ 50 T_H_ cells (FGID, n = 13; iCD, n = 2; cCD, n = 24; UC, n = 14), T_FH_ cells (FGID, n = 19; iCD, n = 5; cCD, n = 27; UC, n = 13), or T_reg_ cells (FGID, n = 20; iCD, n = 5; cCD, n = 28; UC, n = 14) with CDR3β detected. **e**, Mean mTiRP score among PD1Lo and PD1Hi T_FH_ cells. FGID, cCD, and UC diagnostic biopsies with ≥ 50 cells in cell state with CDR3β detected included for analysis (FGID: PD1Lo T_FH_, n = 16; PD1Hi T_FH_, n = 8. iCD: PD1Lo T_FH_, n = 4; PD1Hi T_FH_, n = 3; cCD: PD1Lo T_FH_, n = 15; PD1Hi T_FH_, n = 7. UC: PD1Lo T_FH_, n = 5; PD1Hi T_FH_, n = 9). **f**, Comparison of singlet (CDR3αβ observed only once within a biopsy) and expanded (CDR3αβ observed more than once within a biopsy) PD1Hi T_FH_ mTiRP scores, within FGID, cCD, and UC cells (FGID, 945 singlets, 85 expanded; iCD, 272 singlets, 65 expanded; cCD, 1113 singlets, 127 expanded; UC, 2009 singlets, 250 expanded). T-test used for statistical comparisons. **g, h**, Frequency of TLS-like B cells (GC-like B cells and proliferating B cells) and IgG_1_ plasma cells compared to mean PD1Hi T_FH_ mTiRP. FGID, cCD, and UC diagnostic biopsies included for linear regression analysis contained ≥ 50 PD1Hi T_FH_ cells with CDR3β detected and ≥ 10 TLS-like B cells or IgG_1_ plasma cells (TLS-like B cell regression: FGID, n = 8; cCD, n = 9; UC, n = 10. IgG_1_ regression: FGID, n = 8; cCD, n =8; UC, n = 10). iCD excluded from analysis because only 3 biopsies met criteria. **i**, Regression between mTiRP in PD1Hi T_FH_ cells and canonical T_H_17 gene expression in T_H_17 cells among diagnostic or follow-up UC biopsies with ≥ 50 PD1Hi T_FH_ cells with CDR3β detected and ≥ 10 T_H_17 cells (n = 10). **j**, Comparison of PD1Hi T_FH_ mTiRP score among diagnostic and follow-up UC not-on-anti-TNF (NOA), partial responder (PR) and nonresponder (NR) biopsies with ≥ 50 CDR3β detected (NOA: diagnostic biopsy, n = 4; follow-up biopsy, n = 0. PR: diagnostic biopsy, n = 3; follow-up biopsy, n = 0. NR: diagnostic biopsy, n = 2; follow-up biopsy, n = 1). Kruskal-Wallis test used to compare anti-TNF response groups. Two-tailed Mann-Whitney U test used for grouped comparisons unless otherwise specified. All error bars denote standard error. Shaded areas of regressions show 95% confidence interval.

Our analysis demonstrated that UC biopsies exhibited increased mTiRP scores within CD4 T cells overall, and within T_H_ cells, T_reg_ cells, and T_FH_ cells (**Fig. 7c-d**). Moreover, mTiRP scores were further increased in UC PD1Hi T_FH_ cells relative to PD1Lo T_FH_ cells, and in clonally expanded UC PD1Hi T_FH_ cells relative to singlets, suggesting that more active T_FH_ populations in UC bear increased features of polyreactivity (**Fig. 7e-f**). PD1Hi T_FH_ mTiRP scores were significantly associated with TLS-like B cell (R^2^ = 0.53, p = 0.011) and IgG_1_ plasma cell (R^2^ = 0.65, p = 0.003) frequencies in UC, but not in FGID or cCD (**Fig. 7g-h**), potentially suggesting this TCR phenotype helps drive germinal center reactions in UC.

Considering the overlapping and opposing T_H_17 and PD1Hi T_FH_ phenotypes observed in UC, we examined whether PD1Hi T_FH_ mTiRP would have any relationship to T_H_17 gene expression. Among canonical T_H_17 cytokines, we identified a strong inverse association between UC PD1Hi T_FH_ mTiRP score and UC T_H_17 *IL17A* expression (R^2^ = 0.87, p < 0.001) (**Fig. 7i**). We note that UC NR pre-treatment (and post-treatment) biopsies bore the lowest average PD1Hi T_FH_ mTiRP score among UC participants, further suggesting orthogonal mechanisms of pathology (**Fig. 7j**).

Taken together, these data demonstrate that UC T_FH_ cells display a distinct phenotype from T_FH_ cells in both inflammatory (cCD) and non-inflammatory (FGID) study participants. These phenotypes are associated with polyreactivity and correlate strongly with UC-specific increases in the development of humoral immune activation. However, they are inversely correlated with the T_H_17 features associated with anti-TNF nonresponse, suggesting non-overlapping mechanisms of immune pathology in UC.

#### Autoantibody detection through human proteome array analysis reveals high intra-group variability in pediatric IBD patients

In previous studies, in addition to germinal center activation, adult patients with UC have demonstrated enhanced autoantibody production along with other signs of humoral immune pathogenesis^26,27^. Because of this, we performed a proteome-wide screen of plasma from PREDICT participants to determine the burden of autoantibodies on a per-patient basis. To accomplish this, we used the HuProt microarray (CDI Labs)^80–83^, which assesses immunoglobulins for binding to full-length recombinant human proteins produced in yeast, with coverage against approximately 80% of the human proteome^80–83^. For comparison, we included samples (N=5) from patients diagnosed with autoimmune polyendocrinopathy-candidiasis-ectodermal dystrophy (APECED), a rare condition caused by mutations in *AIRE* that is characterized by a broad range of autoantibodies, which can be considered positive controls^84^. As shown in **Supplemental Fig. 9a** and **Supplemental Table 13**, compared to APECED controls, the most striking finding in PREDICT patients was the high intra-group variability that was evident when comparing the median number of autoantigen hits per patient in each group: patients were present in all PREDICT cohorts with low and high median numbers of autoantigen hits. Overall, the median (interquartile range [IQR]) number of IgG hits was 185.5 (135.75-284.75) for FGID, 244 (124.5-361.5) for iCD, 301.5 (167-391) for cCD, and 215 (127-322) for UC, with each cohort containing patients demonstrating similar numbers of hits compared to the APECED controls (379 [364-437]). Similar variability was observed among IgA autoantigens (**Supplemental Table 13**). While many factors may contribute to the high patient-to-patient variability observed in PREDICT, some of this variability was likely explained by differences in antigen presentation driven by specific HLA:TCR interactions. An example of this, wherein 2 UC patients expressing the HLA-DRB1*15:01 allele developed a distinct pattern of autoantigens, is described below.

#### Patients with the HLA-DRB1*15:01 UC risk allele illustrate the T_FH_:T_H_17 inverse relationship and a distinct autoantibody signature

Given the association of TCR characteristics and UC phenotypes, together with the pronounced inter-patient variability in autoantibody burden we discovered with HuProt analysis, we investigated whether HLA antigen presentation may influence the T_FH_/T_H_17 dichotomy, as well as the presence of patient-specific auto-antibody patterns, that we observed in UC. To accomplish this, we analyzed genomic data from PREDICT UC participants to map their HLA alleles, an analysis which detected several previously-identified HLA-DRB1 alleles associated with UC risk (**Supplemental Fig. 9b**, **Supplemental Tables 1, 12**)^85^. HLA-DRB1*15:01, classically associated with multiple sclerosis, but also identified as a risk allele in UC, was present in two UC participants (participant IDs 001-036 and 001-038, **Supplemental Fig. 9b**)^85,86^. Evaluation of TCR phenotypes revealed that biopsies from these two individuals had the highest PD1Hi T_FH_ mTiRP scores (**Supplemental Fig. 9c**). We also found substantially decreased frequency of T_H_17 cells and decreased expression of T_H_17 *IL17A* in these patients, relative to UC participants without the HLA-DRB1*15:01 allele (**Supplemental Fig. 9d-e**), consistent with the dichotomous relationship between T_H_17- and T_FH_ biology we previously documented in UC (**Fig. 6i-k, 7i**). As predicted, PD1Hi T_FH_ frequency and *CXCL13* expression in both T_H_17 cells and PD1Hi T_FH_ cells were increased in these individuals (**Supplemental Fig. 9d**, **f**). IgG_1_ plasma cells were also increased in these individuals, while IgA_2_ plasma cells were decreased (**Supplemental Fig. 9g**). Interestingly, the “PI3+ Epi.” cell state, an inflamed epithelial cell state likely reflecting microbial interaction, was also increased in the individuals with the HLA-DRB1*15:01 allele (**Supplemental Fig. 9h**).

Further analysis of the HuProt microarray data demonstrated that these participants (IDs 001-036 and 001-038) were jointly reactive to multiple antigens, some of which were uniquely present in this pair. These include autoantibodies directed against ENO1, an autoantigen previously identified in multiple autoimmune disorders^87–91^, as well as two novel targets, YWHAQ^92^, MED18^93^ (**Supplemental Fig. 9i-j**). Together, these data are consistent with a model wherein the combination of HLA type, antigen presentation, and T cell biology contribute to the unique autoantibody profiles observed in UC patients and may further explain the intra-group autoantibody heterogeneity observed across PREDICT.

#### Three axes of immunopathology in pediatric IBD

The combined scRNA-Seq, scTCR-Seq, IHC, and autoantibody analysis have led to a concerted model of pediatric IBD that encompasses three axes of immunopathology, with prominent features associated with T_H_1, T_H_17, and T_FH_ cells (**Supplemental Fig. 10a**, **Supplemental Table 14**) differentially represented across IBD endotypes. While cCD was primarily enriched in T_H_1 features with a moderate increase in T_H_17 features compared to FGID, UC was strongly enriched in T_H_17 and T_FH_ features compared to both patient groups (**Supplemental Fig. 10b**). Although we observed no baseline differences in cCD patients who were eventually treated with anti-TNF that would predict their anti-TNF response status, analysis of post-treatment biopsies identified persistent T_H_17 immunopathology as a signature of less-responsive cCD patients (**Supplemental Fig. 10c-d**). T_H_17 cells were further implicated in anti-TNF response when we examined pre-treatment UC biopsies, which demonstrated an increase in T_H_17 features among NR compared to PR patients (there were no FR to anti-TNF in the PREDICT UC cohort) (**Supplemental Fig. 10e**). Paradoxically, although both T_FH_ and T_H_17 features were enriched in UC, we found orthogonal skewing of these factors: patients with greater enrichment for T_FH_ features had decreased association with T_H_17 features (**Supplemental Fig. 10f**)^87–91^. Taken together, these data provide evidence for multiple disease endotypes within classically defined IBD that can be distinguished by their guiding immunopathologic signatures.

## DISCUSSION

Here we present a multiomic examination of pediatric IBD from patients enrolled in the PREDICT study, identifying the distinguishing cellular and molecular features of cCD and UC, and their response to treatment with anti-TNF therapies. Our in-depth profiling provides a comprehensive cellular and molecular landscape of parenchymal, mesenchymal, and immune cells in the pediatric IBD colon prior to biologic or non-biologic treatment, with critical contextualization provided by non-colon-involved CD, and by pediatric FGID patients, who have GI symptoms, but lack IBD-associated inflammation. Most cellular profiling of adult or pediatric IBD has focused on either CD or UC, without direct comparison between the two disease states ^8,9,30,32,94–97^. Although these studies have been crucial for CD and UC characterization, the lack of direct comparison within the same study prevents the elucidation of critical nuances both between and within the two diseases. The PREDICT data underscore the importance of these direct comparisons, given that, while there were multiple immune states that were dysregulated in both CD and UC compared to FGID, close examination revealed distinct patterns of pathology between these two subtypes, revealing cell states and immune pathways specifically enriched in CD or UC. Previously published studies that have directly compared single-cell transcriptomic profiles of CD and UC have been of much smaller scale (often comparing 6 or fewer patients in one or both groups), lack contextualization with non-inflamed contemporaneously-enrolled controls, and lack a fully treatment-naïve group^31,98–100^, which would limit the disease-specific inferences that could be drawn.

This work is most complementary to a recent study of transcriptomic differences in biologic-naïve (but pre-treated) adults with CD and UC, which utilized 3’ scRNA-Seq to describe 109 cell states with similar phenotypes associated with either CD (including expansion of T_H_1 cells and robust interferon stimulation signatures), or UC (including IgG plasma cells, *CXCL13*+ T_FH_/T_PH_ cells, and T_H_17 cells)^3^. The similarity of these overarching features suggests the pathogenic mechanisms governing the generation of these cell states are preserved between pediatric and adult populations. Our study has additionally enabled an in-depth analysis of T cell plasticity (providing evidence for ex-T_H_17 cells comprising the UC T_H_1 cell state, as well as clonal proliferation of CD8+ T_H_17 cells in UC), TCR phenotypes (demonstrating increased polyreactivity in UC T_FH_ cells), and T_FH_ vs. T_H_17 dynamics (identifying an inverse correlation between T_FH_ features and T_H_17 features in UC), highlighting the central role T cells play in delineating IBD subtypes and treatment response paradigms.

PREDICT demonstrated a profound reduction of T_H_1 cells and T_H_1/*IFNG*-associated cell states upon anti-TNF treatment, consistent with the hypothesis that T cell abrogation is a primary mechanism of anti-TNF efficacy^101–103^. *In vitro* attempts to validate this hypothesis have further suggested that anti-TNF’s ability to prevent TNFRII pro-survival signaling, likely from myeloid cells, is a primary mechanism of drug efficacy^104,105^. Our results are consistent with a model whereby anti-TNF limits T_H_1 differentiation, proliferation, or survival, reducing pathologic IFNγ in the inflamed tissue and contributing to a shift away from inflammatory T cell, NK cell, and macrophage populations. Whereas anti-TNF successfully abrogates T_H_1-mediated inflammation, our results suggest that it is less efficacious at correcting T_H_17-associated pathology^105^. In PREDICT, *IL17A* gene expression was associated with lack of response to anti-TNF, either prior to treatment (in UC), or due to a shift in T cell phenotype following treatment (in cCD). Previous studies have identified a transition to increased T_H_17 features, including increased *IL17A* expression, in treatment-refractory adult CD^105,106^.

Given these results, it is, perhaps, counterintuitive that selected case studies have reported new onset IBD following anti-IL-17A treatment of other rheumatological diseases, and that the only IL-17A cytokine blockade trial in IBD was halted due to adverse events in CD patients receiving the anti-IL17A treatment^107–112^. Moreover, due to IL-17’s other roles in host defense and tissue remodeling, it is likely that this cytokine contributes to both persistence of inflammation and resolution of injury^36,113^. Rather than targeting IL-17 directly, our data suggest that preventing T_H_17 differentiation, for example via JAK inhibition or IL-12/IL-23 blockade, may be more effective in treatment-naïve IBD patients who exhibit greater T_H_17 features, inclusive of *IL17A* expression. This is further reinforced by the emerging use of JAK and IL-12/IL-23 inhibition in IBD treatment, including in those who have failed anti-TNF therapy^72,114–118^. Additionally, neutrophils have also been previously linked to anti-TNF nonresponse^12^. In PREDICT, IHC data demonstrated that neutrophil density was significantly increased in UC. Preventing neutrophil recruitment or activation, for example through IL-1 inhibition, may also be a therapeutic strategy for these patients^12^.

Our analysis provides a framework for combining gene expression and TCR analysis to identify distinct, cell state-influenced TCR phenotypes in autoimmune diseases. Although UC CD4 T cells overall demonstrated an increased disposition towards polyreactive TCR features (described primarily through mTiRP^78^), this signature was particularly enriched in UC PD1Hi T_FH_ cells. This could represent increased reactivity to self-antigens, or potentially to microbial antigens, such as those of the commensal microbiome^78,79^. Additionally, the association of these TCR phenotypes with TLS-like B cells and IgG_1_ plasma cell frequencies may suggest that lymphoid aggregate formation in UC is due to UC-specific antigenic targets in the tissue. Previous studies have suggested that antibodies in UC primarily target commensal antigens, although some targeting of pathogenic bacteria has been observed as well^27,119,120^.

Our evaluation of polyreactive TCR features in T_FH_ highlighted the opposing T_H_17 and T_FH_ biology within UC, which was consistent with increased germinal center activity in some UC patients. To further evaluate this, we performed a proteome-wide autoantibody screen, to determine if the increased T cell polyreactivity in UC was mirrored by increased antibody-mediated autoreactivity.

Our results demonstrated a complex autoantibody landscape in all PRECICT patients, with substantial inter-patient variability being the most prominent feature of the data, and with examples of patients with high autoantibodies in FGID, CD and UC. These data are consistent with multiple factors likely influencing the largely patient-specific autoantibody profile that was measured in PREDICT, including HLA genotype, variations in dietary and microbial exposures, and robustness of humoral immune response. The influence of HLA genotype on the autoantibody profile was highlighted by an evaluation of two UC patients who both expressed the HLA-DRB1*15:01 allele, which has been associated with T cell autoreactivity and cross-reactivity in multiple autoimmune diseases^86,121–124^. These two UC patients were both characterized by T_FH_ features rather than T_H_17 features, and both developed autoantibodies to several antigens not present in any other PREDICT patient, including to ENO1 (a known autoantigen in autoimmune diseases including Hashimoto’s encephalopathy, asthma, juvenile idiopathic arthritis, and Behçet’s disease)^87–91^. These patients highlight the (often unmeasured) influence that HLA genetics can have on the propensity for autoantibody production, underscoring the complex integration of multiple factors in IBD patients that ultimately produce each patient’s unique clinical presentation.

Intriguingly, an experimental murine model of periodontal disease, which is dominated by T_H_17 immunopathology, demonstrated that T_H_17-to-T_FH_ plasticity was critical for generating IgG, restricting growth of oral commensals, and decreasing pathologic tissue damage^125^. Given the similarities to the findings in the PREDICT study, we hypothesize that polyreactive TCR phenotypes may predispose T cells to T_H_17-to-T_FH_ plasticity that could limit T_H_17-mediated colonic inflammation and prevent nonresponse to anti-TNF therapy. While further work is required to address the relationship between T_H_17 and T_FH_ cell biology, it is interesting to note that, in contrast to recent adult UC cohorts which revealed a dominant T_PH_ cell signature together with plasma cell infiltration in UC, pediatric UC harbors a co-existence of T_H_17 and T_FH_ cells of related clonotypes. Thus, this careful study of pediatric participants may be helping to reveal an earlier “transition state” in this disease prior to the formation of more organized lymphoid aggregates.

In summary, our results provide novel insights into the overlapping and distinct biology of pediatric cCD and UC, identifying three prominent axes of immunopathology (T_H_1, T_H_17, and T_FH_) whose interactions and relative magnitude define these two IBD subtypes. They delineate which immune pathways portend suboptimal response to anti-TNF therapy and nominate data-driven approaches to IBD treatment.

## Supporting information

Supplemental Table 1 - Participant and Sample Details

Supplemental Table 2 - UMAP Coordinates

Supplemental Table 3 - Cell State Differential Expression and Markers

Supplemental Table 4 - Milo Differential Abundance

Supplemental Table 5 - IHC Density and scRNA-seq Cell State Frequency

Supplemental Table 6 - Pseudobulk Differential Expression and Gene Set Enrichment

Supplemental Table 7 - Log-Normalized Gene Expression

Supplemental Table 8 - TCR Sequences and Phenotypic Analysis

Supplemental Table 9 - Cell State Frequency Correlations

Supplemental Table 10 - Linear Regressions

Supplemental Table 11 - Gene Set Scoring

Supplemental Table 12 - HLA Alleles

Supplemental Table 13 - HuProt Proteome Microarray

Supplemental Table 14 - Summary of Key Data

## Data Availability

All data produced in the present study are available upon reasonable request to the authors.

## ACKNOWLEDGEMENTS

The authors would like to thank Rachid Hamid for leading study and sample operations at Regeneron for this study, Jason Mighty for operational support for the genomic analyses, Baijun Kou for pathology support, and Mehrdad Rajaei for contributions to development of immunohistochemistry process for this study. J.O.M is a New York Stem Cell Foundation - Robertson Investigator. J.O.M was supported by the AbbVie-Harvard Medical School Alliance, the AGA Research Foundation’s AGA-Takeda Pharmaceuticals Research Scholar Award in IBD - AGA2020-13-01, the Leona M. and Harry B. Helmsley Charitable Trust, The Pew Charitable Trusts Biomedical Scholars, The Broad Next Generation Award, The Chan Zuckerberg Initiative Pediatric Networks, The Mathers Foundation, The Kenneth Rainin Foundation, The Massachusetts Consortium for Pathogen Readiness, The New York Stem Cell Foundation, NIH R01 HL162642, NIH R01 DE031928, NIH P30 DK034854, the NIH CIDER Network, Project NextGen, and The Cell Discovery Network, a collaborative funded by The Manton Foundation and The Warren Alpert Foundation at Boston Children’s Hospital. L.S.K is supported by NIH P01 1P01HL158504, R01 5R01HL095791, U19 U19AI051731, and by the Helmsley Charitable Trust. A.K.S. was supported, in part, by the Searle Scholars Program, the Beckman Young Investigator Program, a Sloan Fellowship in Chemistry, and the NIH (5U24AI118672, 2R01HL095791). L.N. was supported by the National Institute of General Medical Sciences (T32GM007753 and T32GM144273). The research reported is solely the responsibility of the authors and does not necessarily represent the official views of the National Institute of General Medical Sciences or the National Institutes of Health. V.N. was supported by the International mobility of research, technical and administrative staff of research organizations (CZ.02.2.69/0.0/0.0/18_053/0016981). S.B.S. is supported by NIH grants P30 DK034854 and RC2 DK122532, and the Helmsley Charitable Trust. All figures created in *BioRender*.

## METHODS

### Participants and sample collection

#### Study recruitment and clinical parameters

Pediatric patients were enrolled in the Precision Diagnostics in Inflammatory Bowel Disease, Cellular Therapies, and Transplantation (PREDICT) trial (clinicaltrials.gov #NCT03369353), as described previously^9^. Inclusion criteria required participants to be 6 years or older, weigh 10 kg or greater, and to be undergoing evaluation for new diagnosis of inflammatory bowel disease (IBD) or functional gastrointestinal disorder (FGID). Enrollment took place in accordance with Fred Hutchinson Cancer Center Institutional Review Board (Protocol #9730, ethical approval given) and Boston Children’s Hospital Institutional Review Board (Protocol IRB-P00030890, ethical approval given) approved protocols, with written informed consent and assent when applicable. Enrollment for the trial began on May 1, 2017. For this analysis, only enrollees who underwent diagnostic endoscopy prior to February 1, 2022 were included. Diagnoses of FGID, Crohn’s disease (CD), and ulcerative colitis (UC) were made by the patient’s clinician based on endoscopic, histopathologic, and clinical features. Patients were designated as ileal-only Crohn’s disease (iCD) if inflammation was present in the terminal ileum and not in any section of the colon. Designation of colon-involved Crohn’s disease (cCD) required inflammation anywhere in the large intestine or in perianal area. Patients were excluded from the analysis if diagnosed with indeterminate IBD (2 cases), inflammation due to infectious etiology (1 case), or a non-FGID disorder (1 case revised to neurogenic bowel dysfunction). Clinical variables were recorded from enrollment through 2 years post-diagnostic endoscopy. Clinical variables recorded included age, sex, race, disease location and phenotype (using Montreal Criteria), clinical disease severity (calculated using Pediatric Crohn’s Disease Activity Index (PCDAI) or Pediatric Ulcerative Colitis Activity Index (PUCAI) for CD and UC, respectively), anti-TNF use, and anti-TNF response category.

For patients administered anti-TNF (5 iCD patients, 27 cCD patients, 8 UC patients), therapy was initiated within 90 days of diagnostic endoscopy. IBD patients who were not administered anti-TNF within this window were denoted as not on anti-TNF (NOA). Anti-TNF response status was determined at 2 years after diagnostic endoscopy. Full response (FR) was defined as clinical symptom control and biochemical response with a PCDAI score less than 12.5 or PUCAI score less than 10 on maintenance anti-TNF therapy. Partial response (PR) was defined as a lack of complete clinical symptom control and biochemical response with documented dose escalation of anti-TNF therapy and a possible change to a new therapeutic. Nonresponse (NR) was defined as complete lack of clinical and biochemical response to anti-TNF therapy.

#### Colon and blood collection

Colon and whole blood biopsies were obtained during the diagnostic endoscopy visit. Whole blood was drawn into EDTA anticoagulant and frozen at −80° C until required for genomic DNA experiments. Per protocol, up to 2 pinch biopsies were taken from non-necrotic areas of the colon, with preference for areas with marked erythema or edema. If no inflammation was observed, biopsies were obtained from left colon by default. Pinch biopsies were immediately placed into RPMI 1640 (ThermoFisher, 21870-076) on ice before subsequent processing.

### Experimental methods

#### Pinch biopsy processing

Pinch biopsy processing was performed using a modified version of a previously published protocol^9,126^. While intact, biopsy pinches were handled using a P1000 pipette applying gentle suction, and all centrifugation steps done in a temperature-controlled 4°C centrifuge. Biopsy pinches were first rinsed in 25 mL PBS (ThermoFisher 14190-144) and allowed to settle. Each individual pinch was then transferred to 10 mL epithelial cell solution (ECS) (HBSS Ca/Mg-Free [ThermoFisher 14175-103], 100 U/mL penicillin [ThermoFisher 15140-122], 100 μg/mL streptomycin [ThermoFisher 15140-122], 10 mM HEPES [ThermoFisher 15630-080], and 2% FBS [ThermoFisher SH3007103HI]), freshly supplemented with 10 mM EDTA [ThermoFisher AM9261]. Separation of the epithelial layer (EPI) from the underlying lamina propria (LP) was performed for 15 minutes at 37°C with rotation at 700 RPM. The tube was then removed and placed on ice immediately for 10 minutes before shaking vigorously 20 times and undergoing 10 seconds on high vortex. Visual macroscopic inspection of the tube at this point yielded visible epithelial sheets.

The LP tissue fraction was carefully removed and placed into a large volume (45 mL) of ice-cold PBS to rinse before transferring to 10 mL of enzymatic digestion mix (Base: RPMI1640 [ThermoFisher 21870-076], 100 U/ml penicillin [ThermoFisher 15140-122], 100 μg/mL streptomycin [ThermoFisher 15140-122], 10 mM HEPES [ThermoFisher 15630-080], 2% FBS [ThermoFisher SH3007103HI], and 50 μg/mL gentamicin [ThermoFisher 15750-060]), freshly supplemented immediately prior with 100 μg/mL of Liberase TM [Roche 5401127001] and 100 μg/mL of DNase I [Sigma D5025-150KU]), at 37°C with rotation at 700 rpm for 30 minutes. LP enzymatic dissociation was quenched by addition 80 μL of 0.5M EDTA and placed on ice for five minutes. Samples were typically fully dissociated at this step and after gentle trituration with a P1000 pipette filtered through a 40 μm cell strainer (VWR 21008-949) into a new 50 mL conical tube and rinsed with ice-cold PBS to 35 mL total volume. This tube was spun down at 500g for 10 minutes and resuspended in 1.2 mL ECS. The tube was then spun down at 800g for 2 minutes and resuspended in 500 μL of ACK lysis buffer (Gibco A10492-01) and placed on ice for 3 minutes. 500 μL 0.2% FBS (ThermoFisher SH3007103HI) in DPBS (ThermoFisher 14-190-144) was added to post-lysis solution before centrifugation at 800g for 2 minutes and resuspension in 700 μL ice-cold PBS with 0.4% BSA (ThermoFisher AM2616). LP cells were filtered through a 40 μm strainer then rinsed with an additional 700 μL ice-cold PBS with 0.4% BSA. LP cells were centrifuged at 500g for 5 minutes, then resuspended in 60 μL PBS + 0.4% BSA.

During LP processing, ECS was added to EPI-containing tube to 35 reach mL before centrifugation at 600g for 10 minutes and resuspension in 1 mL ECS. Cells were transferred to a 1.5mL Eppendorf tube pre-coated with 0.4% BSA in PBS in order to minimize time spent centrifuging and obtain a more concentrated cell pellet. Cells were spun down at 800g for 3 minutes, resuspended with 1 mL ECS, and centrifuged again at 800g for 3 minutes. Pellets were resuspended in ACK lysis buffer [ThermoFisher A1049201] for 2 minutes on ice to remove red blood cells, even if no RBC contamination was visibly observed, in order to maintain consistency across samples. 500 μL ECS without FBS was added to EPI cells before centrifugation at 800g for 5 minutes. Cells were washed (resuspended in 1 mL ECS without FBS, centrifuged at 800g for 3 minutes) and resuspended in 1 mL 37° C TrypLE express enzyme [ThermoFisher 12604-013]. After adequate mixing, another 400 μL TrypLE solution was added before additional mixing. Cells were incubated in TrypLE for 4 minutes in a 37°C bath followed by gentle trituration with a P1000 pipette. Cells were incubated for another 3 minutes in a 37°C bath followed by gentle trituration, and an additional 3 minute incubation at 37° C. Cells were centrifuged at 800g for 3 minutes then resuspended in 700 μL PBS + 0.4% BSA. Resuspended cells were filtered through 40 μm strainer and rinsed with 700 μL PBS + 0.4% BSA before being transferred to a new BSA-coated tube. Cells were centrifuged at 500g for 5 minutes and resuspended in 50 μL PBS + 0.4% BSA. Cells from both EPI and LP fractions were counted and prepared as a single-cell suspension for scRNA-seq.

#### Single-cell RNA sequencing, read alignment, and ambient mRNA correction

Single cells were loaded onto 5’ library chips as per the manufacturer’s protocol for Chromium Single Cell 5’ Library and Gel Bead Kit (10X Genomics). Biopsies were processed with 5’ v1 kits November 2020 and prior (Chromium Single Cell 5ʹ Library and Gel Bead Kit, PN-1000006; Chromium Single Cell V(D)J Enrichment Kit, Human T Cell, PN-1000005), or 5’ v2 kits (Chromium Next GEM Single Cell 5ʹ v2 Library and Gel Bead Kit, PN-1000263; Chromium Single Cell V(D)J Enrichment Kit, Human T Cell, PN-1000252) since November 2020. The EPI and LP fractions were processed in separate channels of the 10X Chromium Single Cell Platform. An input of 20,000 single cells was added to each channel. Briefly, single cells were portioned into Gel Beads in Emulsion (GEMs) in the Chromium controller with cell lysis and barcoded reverse transcription of RNA, followed by cDNA amplification, enzymatic fragmentation and 5’ adaptor and sample index attachment.

For single cell libraries prepared using the 5’ v2 assay from 10x Genomics, paired-end sequencing was performed on Illumina NovaSeq 6000 for RNA-seq libraries (Read 1 26-bp for UMI and cell barcode, Read 2 80-bp for transcript read, with 10-bp i7 and 10-bp i5 reads) and for V(D)J libraries (Read 1 150-bp, 10-bp i7, 10-bp i5, Read 2 150-bp). For single cell libraries prepared using the 5’ v1 assay from 10x Genomics, paired-end sequencing was performed on Illumina NovaSeq 6000 for RNA-seq libraries (Read 1 26-bp for UMI and cell barcode, Read 2 80-bp for transcript read, with 8-bp i7 and 0-bp i5 reads) and for V(D)J libraries (Read 1 150-bp, 8-bp i7, 0-bp i5, Read 2 150-bp). Quality-filtered base calls were converted to demultiplexed FASTQ files. For all RNA-seq libraries, Cell Ranger Single Cell Software Suite (10X Genomics, 7.2.0) was used to perform transcriptome alignment, filtering, and UMI counting. The human GRCh38-1.2.0 genome assembly was used for the alignment. Intronic reads were not included in alignment. For all V(D)J libraries, Cell Ranger Single Cell Software Suite (10X Genomics, 7.2.0) was used to perform de novo assembly of read pairs into contigs followed by alignment and annotations of contigs against the germline segment V(D)J reference sequences from the GRCh38-alts-ensembl-2.0.0 genome assembly.

For all aligned RNA-seq libraries, CellBender “remove-background” was used to reduce contaminating signal due to ambient mRNA^127,128^. Raw Cell Ranger-aligned counts matrix were used as input. Default remove-background settings were used for all samples, with exception of learning rate (set to 3×10^-5^ for all samples, instead of default 10×10^-5^)

#### scRNA-seq data processing

Cell x gene matrix outputs from CellBender were aggregated into a single object using Seurat (version 5.0)^129^. Samples were converted to AnnData format using zellkonverter^130^. The AnnData object was processed with scanpy log-normalization, variable gene selection (flavor = seurat_v3, batch = 10x version, n_top_genes = 3000), PCA dimensional reduction, and coarse leiden clustering (n_neighbors = 20, n_pcs = 20, leiden resolution = 0.1)^10^. Coarse clusters were grouped into one of 6 groups: Colonic Epithelium (ColEpi); Stromal and Endothelial Cells (Stroma); Myeloid Cells (Myeloid); Plasma Cells (Plasma); B Cells (BCell); and T Cells, NK Cells, and Innate Lymphoid Cells (TNKILC). Clusters that were low-quality based on mitochondrial gene expression, number of features detected, number of UMIs detected, or a combination of these metrics, were not included in any of the above groups and discarded from further processing. Sample-specific clusters were also not included.

After partitioning into coarse cell types, each cell type was log-normalized and underwent 3 rounds of scVI model training, clustering, differential expression, annotation, and low-quality cell removal. For each round, an scVI model was trained and the scVI latent space was used for dimensional reduction, and between-cluster differential expression performed within the scVI framework^131^. Settings for scVI: n_layers = 2, n_latent = 30, n_hidden = 128, dropout_rate = 0.1, gene_likelihood = “nb”, max_epochs = 250 (with early stopping), batch_key = “version_10x”.

AnnData objects were reduced to top n variable genes prior to scVI model training. Number of variable gene selected differed by cellgroup (number of genes used for scVI input in final round of annotation and QC: ColEpi = 2000; Stroma = 5000; Myeloid = 5000; Plasma = 750; BCell = 3000; TNKILC = 5000). For Plasma and BCell types, additional gene filtering was performed to exclude variable immunoglobulin genes (pattern matching for “^IG[HKL][VJC]”). This step was taken to reduce the ability of variable immune receptor genes to dominate model fitting and latent space generation in plasma cells/plasmablasts (constant genes were retained). Additional genes matching the following patterns were removed from scVI training as they appeared to dominate clustering without clear biological function: “^CH17’”, “^RP11”, “^IGLL”, “^AC[0-9]{5}”. Conversely, genes matching the following pattern were explicitly included as input (if not already an highly variable gene) for plasma cell scVI model training, to ensure constant genes would be included: “IGH[ADEGM][1-4]*$”

For cluster annotation, annotation was based on marker genes identified through literature review. Annotations were defined at additional levels within each cell type (level 1): cell subtype (level 2) and cell state (level 3). If appropriate and supported by literature, clusters were recombined into overarching cell types to obtain “level 2 annotations.” Conversely, if clusters were observed to be heterogeneous through differential expression analysis and visualization, subclustering was attempted, along with differential expression and annotation to obtain level 3 annotations (“cell state”). Annotations from final round of scVI processing were used for this study.

#### scTCR-seq data processing

After Cell Ranger alignment to the V(D)J reference, the filtered contigs files from each sample were combined into a single data frame. In some cases, more than one alpha chain was detected per barcode. In very rare cases, more than one beta chain was detected per barcode. To simplify data analysis, a single alpha or beta chain was selected for downstream processing. First, barcodes with only a CDR3 listed were excluded (these chains lacked values for CDRs 1 and 2 as well as framework regions 1-4). The selection preference order was: (1) which chain had the most UMIs, (2) which chain had the most reads, (3) which chain is listed first (this condition was rarely reached in the dataset). Cell annotations from the RNA-seq processing step were merged with TCR data through paired barcode sequences.

#### Immunofluorescence Microscopy

A fully automated multiplex immunohistochemistry assay was performed on the Ventana Discovery ULTRA platform (Ventana Medical Systems, Tucson, AZ) as previously described^9,132^. Optimal concentrations of each antibody were determined, and they were applied in the following sequence and detected with the indicated fluorophore.

Pan-Immune Panel 1:

1. Rabbit anti-BCMA (clone E6D7B, Cell Signaling, SC#88183) was detected with DISCOVERY Rhodamine 6G (Roche, Part number: 7988168001).
2. Rabbit anti-ribonuclease 3/ECP (clone EPR20357, Abcam, ab207429) was detected with DISCOVERY DCC (Roche, Part number: 7988192001).
3. Rabbit anti-myeloperoxidase (clone EPR20257, Abcam, ab208670) was detected with DISCOVERY Cy5 (Roche, Part number: 7551215001).
4. Rabbit anti-CD11c (clone EP1347Y, Abcam, ab52623) was detected with DISCOVERY FAM (Roche, Part number: 7988150001).
5. Rabbit anti-NCR1 (clone EPR23097-35, Abcam, ab233558) was detected with Cy7-Goat anti-Rabbit-IgG (H+L) (AAT Bioquest, Cat 16872).
6. Mouse anti-pan keratin (clones AE1/AE3/PCK26, Roche, Part number 5266840001) was detected with DISCOVERY RED 610 (Roche, Part number: 7988176001).

Pan-Immune Panel 2:

1. Rabbit anti-CD20 (clone SP32, Abcam, ab64088) was detected with DISCOVERY Rhodamine 6G (Roche, Part number: 7988168001).
2. Rabbit anti-CD3 (clone SP162, Abcam, ab135372) was detected with DISCOVERY DCC (Roche, Part number: 7988192001).
3. Rabbit anti-CD8 (clone SP239, Abcam, ab178089) was detected with DISCOVERY RED 610 (Roche, Part number: 7988176001).
4. Rabbit anti-CD68 (clone SP251, Abcam, ab192847) was detected with DISCOVERY Cy5 (Roche, Part number: 7551215001).
5. Rabbit anti-FOXP3 (clone SP97, Abcam, ab99963) was detected with DISCOVERY FAM (Roche, Part number: 7988150001).
6. Mouse anti-pan keratin (clones AE1/AE3/PCK26, Roche, Part number 5266840001) was detected with Cy7-Goat anti-Mouse-IgG (H+L) (AAT Bioquest, Cat 16856).

Following staining, the tissue was counter-stained and cover slipped with Invitrogen ProLong Gold Antifade Mountant with NucBlue. Whole slide imaging was performed on the Zeiss Axioscan which was equipped with a Colibri light source.

#### Whole exome sequencing

For analyses of common variants, we used Illumina GSA array genotyping data and performed imputation with the TOPMed imputation server and reference panel^133^. Exome sequencing was performed at the Regeneron Genetics Center using a custom automated sample preparation approach. Samples were captured with IDT xGen v1 probes and sequenced using Illumina HiSeq 2500-v4 or Illumina NovaSeq instruments, with 75-bp paired-end reads and two index reads. The GRCh38 human genome reference sequence and Ensembl version 100 gene definitions were used for variant calling using DeepVariant and for annotation^134^.

#### HLA imputation

We imputed HLA alleles using the Michigan Imputation Server^133^, HLA reference panel multiethnic-hla-panel-4digit-v2@1.0.0^135^. Prior to imputation, in order to maximize the number of informative variants for HLA imputation, the PREDICT IBD sample was merged with n=1,000 random individuals from a Regeneron Genetics Center multi-ethnic cohort genotyped on the same array version. Variants in the MHC region chr6:28-34Mb and present in the HLA reference panel were extracted from TOPMED imputed genotype probabilities and hard-called with certainty threshold 0.9. The resulting dataset was QC-filtered for 12,101 variants with less than 0.1% missing genotype calls and HWE Pval > 1e-10. (All samples had less than 1% missing genotype calls.)

#### Proteome Microarray

The HuProt proteome microarray (CDI Labs, USA) was used to assay plasma for autoreactivity against the human proteome^80–83^. HuProt arrays and plasma samples were diluted with CDI proprietary CDISampleBuffer (CDI Labs, USA) at 1:1000 and incubated with CDIArrayBlock (CDI Labs, USA) at room temperature with gentle shaking for 1 hour. After the blocking period, each sample was probed onto a HuProt microarray at room temperature for 1 hour with gentle shaking. The arrays were then washed with TBS-Tween (1x TBS / 0.1% Tween-20) 3 times for 10 minutes each. Following washes, the arrays were probed with fluorescent anti-human IgA or IgG secondary for 1 hour in the dark with gentle shaking. 3 washes were performed with TBS-Tween for 10 minutes each, followed by 3 rinses with ddH_2_0. The arrays were dried with compressed CO_2_. GenePix 4000B scanner was used to scan arrays. Scanned images were aligned to the array layout. Raw fluorescence intensity values were extracted using GenePix software for further data processing.

### Quantification and statistical analysis

#### Gene set enrichment analysis

Gene set enrichment analysis was performed using fgsea^136^. The DESeq2 “stat” value (Log_2_FC/SE_log2FC_) was used as gene-level statistical input for fgsea. 1000 permutations were used for fgsea calculation of enrichment scores. Reactome gene sets were used as input (version 2023.2, obtained through MSigDB C2 curated gene set)^137,138^. Gene sets were included if they had at least 15 genes and no more than 100 genes.

#### Cell frequency analysis

Per-sample mean cell frequencies were calculated using custom code. Cells were grouped first by their sample or biopsy of origin (relevant grouping listed in text and/or figure captions; “sample” will be used throughout methods), then by the annotation of interest (e.g. CD4 Naïve T cells). The number of cells per sample and annotation was tallied. Unless otherwise stated, the number of cells was divided by a relative grouping, e.g. a cell subtype (e.g. Naïve T cells), a cell type (e.g. T cells), or the entire independent sample (e.g. “Relative to Sample”). For per-biopsy group comparisons, biopsies were only included for analysis if the relative grouping had at least 10 cells. This was an empirical measure taken to prevent statistical conclusions driven by low overall cell detection. Statistical testing was performed with ggpubr’s “stat_compare_means()” function^139^.

#### Quantitative feature analysis

Per-sample quantitative feature (e.g. gene expression, module score, mTiRP, etc.) values were calculated using custom code. Data were aggregated based on sample of origin, then by cellular annotation(s) of interest. Mean values were calculated per combination of sample and cell annotation. For gene expression, log-normalized values were used as input for mean calculation. Samples were required to meet a minimum number of cells within sample and cell annotation combination to be included for analysis. 10 cells were required for calculation of gene-based metrics, and 50 cells were required for TCR-based metrics.

#### Milo differential abundance analysis

Only baseline, non-iCD samples were used as input for analysis with MiloR^13^. Milo analysis was performed on a per cell type basis (ColEpi, Stroma, Myeloid, Plasma, BCell, TNKILC). For each cell type, the data object was downsampled to the number of cells in the smallest disease category between FGID, cCD, and UC. This step was performed to reduce the effect of differing total number of cells skewing differential abundance results. Downsampling was performed on a per disease category basis, irrespective of sample of origin within cell types. After downsampling, any remaining biopsies with fewer than 5 cells were removed before proceeding with analysis. Following biopsy removal step, the Milo object was created and kNN graph generated with the cell group scVI latent space used as the reduced dimension (all 30 scVI dimensions used as input). Because there were many samples in this analysis (∼70), we set k = 200 to ensure the average neighborhood would have cells numbering greater than 5 x number of samples (reducing the likelihood of a single sample comprising a neighborhood), per package maintainer recommendations^13^. We verified that each cell group fulfilled this heuristic, then calculated neighborhood distances and built the neighborhood graph, using Milo functions with default inputs and latent space inputs as described above. Following graph construction, we tested for differential abundance with the scVI latent space used as the reduced dimension and a design formula specifying 10x version as a batch effect: “∼version_10x + disease_group”. Alpha value was set to 0.1 for identifying differentially abundant neighborhoods. Neighborhoods with annotation fraction < 0.7 were excluded from visualization. 12 cell states did not meet the annotation fraction threshold in any neighborhood and were not visualized: Lymph. Endo., IgG4 PC, Prlf. IgG PC, Prlf. IgM PC, IgD PC, IgE PC, Plasmablast, Prlf. Th17.1, Prlf. Tfh, Prlf. Treg, Vg9d2 TC, and NKT.

#### Cell state correlation analysis

Cell state correlation analysis was performed using per-cell type frequencies of annotated cell states. Cell states were excluded if they did not have at least 100 cells (2 cell states excluded:

IgD PC and IgE PC). Only baseline biopsies were included for correlation analysis. The previously described cell frequency calculation method was used to generate per cell type frequencies for each cell state in each biopsy. Pearson correlation was used to compare biopsy frequencies of T_H_1, T_H_17, and PD1hi T_FH_ cells against all cell states excluding IgD PC and IgE PC. Test statistics were generated using “cor.test” function from the R stats package (version 4.3.1). Cell state correlations with a p value < 0.001 were selected as part of T_H_1, T_H_17, or PD1Hi T_FH_ network. Cell states in network were visualized with ggraph (version 2.2.1). Only edges with a correlation coefficient > 0.35 were visualized. Graph layout algorithm set to “kk”, maximum iterations = 150. Node location manually adjusted when necessary for visualization of all cell states.

To provide a quantitative measure of the usage of cell states, we calculated the geometric mean of cell state frequencies, as has been performed previously^30^. Briefly, we took the logarithm value of the associated cell state frequencies in each biopsy, calculated the mean of all cell state frequencies in logarithm space, then exponentiated the average value:

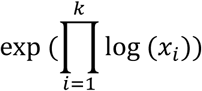

Where *x* is the cell state proportion, *i* is the index of the cell state in the correlation network, and *k* is the number of total cell states in the correlation network. Because calculation of the geometric mean requires a logarithmic transformation, we added a “pseudoproportion” of 1/1000 to any cell state which was not identified in a biopsy.

#### Pseudobulk differential gene expression analysis

For differential expression (DE) analysis, a single-cell object containing aggregated, annotated cells from each cell type was filtered to include only the biopsies of interest for that comparison. Raw counts from each cell type within each biopsy were aggregated into pseudobulk format using the Seurat “AggregateExpression()” function^140^. Gene selection was performed by filtering for genes with ≥ 250 counts in at least as many samples as were in the smaller of the two groups being compared (e.g. if comparing 15 cCD FR and 10 cCD PR samples, genes were included if there were ≥ 250 counts in at least 10 samples overall in the cell type of interest). For the combined myeloid and TNKILC pre-vs. post-treatment analysis, a 500 count minimum was used. All input genes are included in supplemental DE tables. DESeq2 was run with default settings, using UMI data from the pseudobulk object^141^.

#### Gene set scoring

Decoupler python package (version 1.5.0) was used for all gene set scoring^50^. For TNF scoring, the PROGENy database was used to select genes involved in TNF pathway activity^51^. Top 500 genes were obtained initially. The genes were then filtered to only include those with p-value < 1×10^-10^, to increase confidence that the genes included in the network play a role in TNF signaling. This resulted in 225 input genes. These genes were used as input for decoupler’s multivariate linear model (mlm), with weights provided by PROGENy database entries^50,51^. Decoupler was performed on individual cell types (e.g. Myeloid, TNKILC) and mlm estimate values were used for data analysis. For IL-17 pathway scoring, genes that were listed as cell extrinsic and downstream of IL-17RA/IL-17RC signaling in Kyoto Encyclopedia of Genes and Genomes (KEGG) pathway “IL-17 SIGNALING PATHWAY” were investigated for gene expression in this dataset^46^. Genes with robustly detectable expression were retained for further analysis and grouped into KEGG-provided gene groups: Chemokines (*CXCL1, CXCL2, CXCL5, CXCL8, CXCL10, CCL2, CCL7, CCL20*), Cytokines (*IL6, TNF, PTGS2, CSF2, CSF3*), Anti-microbial (*DEFA5, DEFA6, DEFB1, DEFB4A, DEFB4B, DEFB124, MUC5B, S100A7, S100A8, S100A9, LCN2*), Tissue remodeling (*MMP1, MMP3, MMP9, MMP13*). An unweighted network of these genes was created with the gene group as the source and the gene as the target. This network was used as input with each cell type for decoupler’s weighted sum algorithm^50^. However, equal weight was given to all genes in the network. Normalized wsum values were used for data analysis. The same analytical steps as IL-17 gene set scoring were used for T_H_1 and T_H_17 cytokine (*IFNG*, *TNF*, *IL17A*, *IL17F*, *IL21*, *IL22*, *IL26*) scoring.

#### Regression analysis

Comparison of multiple metrics through regression analysis was performed using a combination of custom code and ggpmisc (version 0.5.5)^142^. Cell frequency and feature quantification was performed as described in “Cell frequency analysis” and “Quantitative feature analysis.” The regression analysis was performed using “stat_poly_eq” (for statistics) and “stat_poly_line” (for visualization) functions. The method was set to “lm” for all regressions. A 95% confidence interval used for shading.

#### T Cell Clonal Analysis

A T cell was defined as clonally expanded if its paired CDR3α and CDR3β amino acid sequences were observed in at least one other cell in the same biopsy. The number of cells in a clonotype (paired CDR3α+CDR3β) per biopsy was quantified and reported as the level of clonal expansion: 1 (singlet), 2, 3, 4, or 5+. Cells that did not have a CDR3α or CDR3β were not considered for clonal expansion analysis.

For T_H_-T_FH_ cell clonal overlap analysis, the dataset was reduced to T_H_1, T_H_17, PD1Hi T_FH_, and PD1Lo T_FH_ cell states. Additional filtering was performed to ensure paired CDR3α+CDR3β was present in each cell. Cells were grouped by paired CDR3 per biopsy and the number of cell subtypes within each biopsy+CDR3 pair was determined. Clonotypes spanning more than one cell subtype were considered to have clonal overlap. Visualization of clonal overlap was performed with ggraph (version 2.2.1).

Clonal overlap between Prlf. T_H_17.1 and T_H_1 or T_H_17 was performed in an identical manner, except that cell states were used to define overlap rather than cell subtypes.

#### Fluorescence microscopy quantification

Quantitative image analysis was performed using HALO Indica Labs Hyperplex module (IndicaLabs, Albuquerque, NM). For each sample, images from the pan-immune panels and the pan-CK marker were fused to generate a single image. A classifier was first applied to detect the tissue in each section and an automatic annotation was generated as “whole section.” A second classifier was applied using panCK and DAPI channels to define the epithelium layer (panCK+) and the lamina propria (panCK-). Automated annotations were generated as “epithelium” and as “lamina propria”. Tertiary lymphoid structures (TLS) were annotated by a pathologist. Their number, frequency, size, and cell composition were measured. Immune cells were detected in each area of interest (whole section, epithelium, and lamina propria for pan-immune panel 1; whole section, epithelium, lamina propria, and tertiary lymphoid structures for pan-immune panel 2). T cells were defined as CD3+, B cells as CD20+, myeloid cells as CD68+. For each T cell subset, CD8 T cells were defined as CD3+CD8+FOXP3-, CD4 as CD3+CD8-, Tregs as CD3+CD8-FOXP3+ and CD4 conventional (non-Treg CD4) as CD3+CD8-FOXP3-. Plasma cells were defined as BCMA+. Eosinophils were defined as ECP+. Dendritic cells were defined as CD11c+. Neutrophils were defined as MPO+. NK cells were defined as NCR1+. Numbers of positive cells for each immune subset were counted and their density measured.

#### Pseudotime Analysis

Diffusion pseudotime was calculated using scanpy^10^. For pseudotime analysis, a subset of the TNKILC cell group was created from CD4 T cell states (NaiveCD4, MemoryCD4, Int. T_H_, T_H_1, T_H_17, Prlf. T_H_17.1, PD1Lo T_FH_, PD1Hi T_FH_, Prlf. T_FH_, T_reg_, LEF1+ T_reg_, LAG3+ T_reg_, Prlf. T_reg_, CD4 CTL). Nearest neighbors were recalculated on this subset, with n_neighbors = 20 and all 30 scVI latent dimensions used. The starting position for the pseudotime analysis was selected as the cell with the most positive value in the UMAP2 dimension, which corresponded to a cell in the NaiveCD4 cell state. Diffusion pseudotime was calculated using scanpy’s “tl.dpt” function, with n_branchings = 0 and n_dcs = 10. Gene expression of cells ordered by pseudotime was visualized using scanpy’s “pl.paga_path” function, with “normalize_to_zero_one = True”.

#### CDR3 Metrics

CDR3 length was quantified as the number of amino acids in the CDR3, as determined by Cell Ranger alignment and TCR classification. Previous studies have highlighted that the middle residues of the CDR3β, determined by N and P nucleotides between regions belonging to the V gene and J gene, are most likely to directly participate in contacts between TCRs and antigenic peptides^75^. Thus, we focused our subsequent biophysical analysis on the middle region of the CDR3β. The CDR3β middle region was calculated only for TCRs with CDR3β lengths between 12-17 amino acids, in accordance with definition for T cell regulatory potential (TiRP) score^78^. The Cell Ranger definition of CDR3 includes junctional amino acids at IMGT positions 104 and 118^143^. The CDR3β middle region was defined as all amino acids from the 5^th^ amino acid from the beginning to the 6^th^ from the end, corresponding to P108 and P112 in IMGT numbering^143^. mTiRP score was calculated using publicly available code from Lagattuta et al. (https://github.com/immunogenomics/TiRP), with Cell Ranger-determined V gene and CDR3β used as input^78^. CDR3β middle-region biophysical properties, including hydrophobicity (“grand average of hydrophobicity”, or “gravy”), size, aliphatic propensity, and polarity, were calculated with “alakazam” R package (version 1.3.0). CDR3β middle-region amino acids were used as input to alakazam aminoAcidProperties() function with nucleotides set to FALSE, all other settings default. Normalized z-scores were generated through transformation to standard score within the dataset: 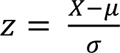 where X is the vector of all metric values, μ is the metric’s mean within the dataset, and σ is the metric’s standard deviation within the dataset. Given the inherent heterogeneity in TCR features, we limited sample-comparison analysis to biopsies with at least 50 CDR3β sequences detected in the cell grouping of interest.

#### Autoantigen analysis

Data from HuProt proteome microarray was processed for autoantigen reactivity among PREDICT participants^80–83^. Briefly, median fluorescence intensity (MFI) of each replicate spot pair on the protein microarray was averaged. Replicates with values differing by two-fold or greater were discarded from further analysis. log_2_(MFI) was calculated to stabilize variance across cohorts. Microarray spots with readings 4-fold above secondary-only median were considered false-positives and removed. Spots that were not detected were also removed. Z-scores were calculated within each sample array for all spots passing QC. Autoantigen detection (binary variable) was defined as spots with readings 4-fold above secondary-only median for the relevant spot, and with a Z-score > +3 within that sample array. Arithmetic mean of values was used for visual representation when an antigen was present more than once on the proteome microarray, and thus had multiple independent readings. An antigen was considered a hit if the antigen met Z-score-based hit criteria in 1 or more positions on the microarray. ComplexHeatmap (version 2.24.0) was used to generate antigen heatmaps and any accompanying dendrogram using default settings^144,145^.

#### T_H_1, T_H_17, and T_FH_ feature summary

Features that were associated with T_H_1 (T_H_1 cell state frequency, geometric mean of T_H_1-associated cell state frequencies, T_H_ *IFNG* expression, myeloid and TNKILC cell type *STAT1* expression, and myeloid and TNKILC TNF pathway score), T_H_17 (T_H_17 cell state frequency, geometric mean of T_H_17-associated cell state frequencies, T_H_ *IL17A* expression, ColEpi antimicrobial gene set score, ColEpi chemokine gene set score), and T_FH_ (PD1Hi T_FH_ cell state frequency, geometric mean of PD1Hi T_FH_-associated cell state frequencies, T_FH_ *CXCL13* expression, IgG_1_ plasma cell state frequency, and T_FH_ mTiRP score) were used as inputs to generate per-biopsy summaries of overall T_H_1, T_H_17, and T_FH_ tendencies. Per-biopsy values for each feature were generated as described for the relevant feature. Each feature was scaled from 0 to 1, corresponding to the minimum and maximum values of that feature within the dataset. The arithmetic mean of each feature within each feature set (T_H_1, T_H_17, or T_FH_) was then averaged per biopsy. The average value of each scaled feature was used to represent biopsies in the per-biopsy radar plot. For group-based radar plots created with ggradar (version 0.2), the arithmetic mean of the scaled feature set among all biopsies within the group was used to represent the group value on the radar plot.

## LIST OF SUPPLEMENTAL TABLES

Supplemental Table 1 - Participant and Sample Details

Supplemental Table 2 - UMAP Coordinates

Supplemental Table 3 - Cell State Differential Expression and Markers

Supplemental Table 4 - Milo Differential Abundance

Supplemental Table 5 - IHC Density and scRNA-seq Cell State Frequency

Supplemental Table 6 - Pseudobulk Differential Expression and Gene Set Enrichment

Supplemental Table 7 - Log-Normalized Gene Expression

Supplemental Table 8 - TCR Sequences and Phenotypic Analysis

Supplemental Table 9 - Cell State Frequency Correlations

Supplemental Table 10 - Linear Regressions

Supplemental Table 11 - Gene Set Scoring

Supplemental Table 12 - HLA Alleles

Supplemental Table 13 - HuProt Proteome Microarray

Supplemental Table 14 - Summary of Key Data

## SUPPLEMENTAL FIGURES

**Supplemental Fig. 1.**
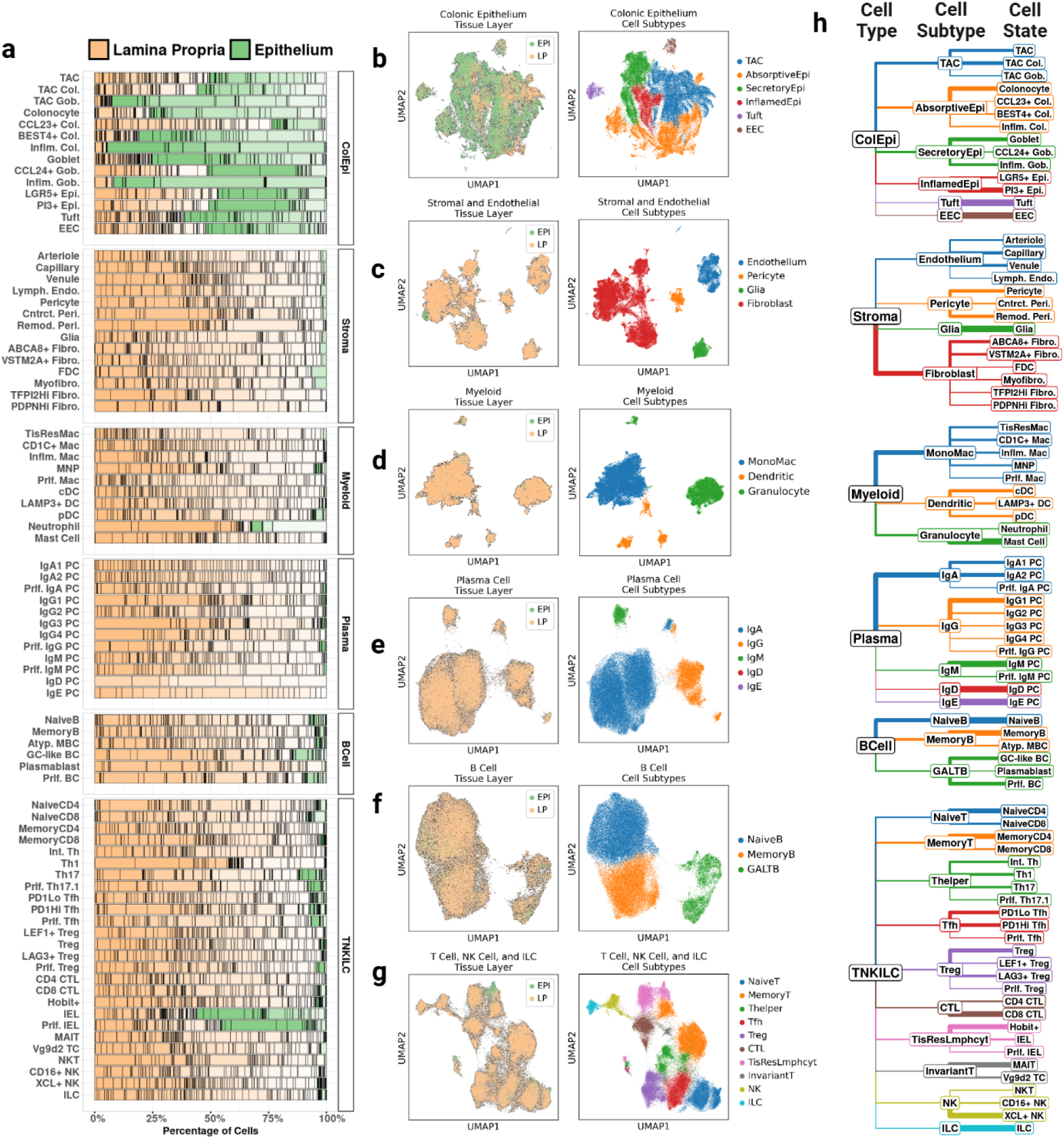
Comprehensive cellular annotation reveals cell state compartmentalization in epithelium or lamina propria **a**, Proportion of annotated cellular states identified within LP (beige) or EPI (green) layer from colonic biopsies. Individual samples are assigned to gradations of layer color, marked with vertical line between distinct samples. **b**, **c**, **d**, **e**, **f**, **g**, UMAPs highlighting cells attributed to EPI or LP samples (left), with corresponding cell subtype annotations for each cell type: colonic epithelium (**b**), stromal and endothelial cells (**c**), myeloid cells (**d**), plasma cells (**e**), B cells (**f**), and T, NK, and ILCs (**g**). (**h**) Visual tree of each cell state, relative to its corresponding cell subtype and cell type. Branch colors match the cell subtype annotation colors in **b**-**g**. Branch width corresponds to the proportion of cells in child node relative to parent (cell subtype/cell type; cell state/cell subtype). UMAP, uniform manifold approximation and projection. LP, lamina propria. EPI, epithelium. NK, natural killer. ILC, innate lymphoid cell.

**Supplemental Fig. 2.**
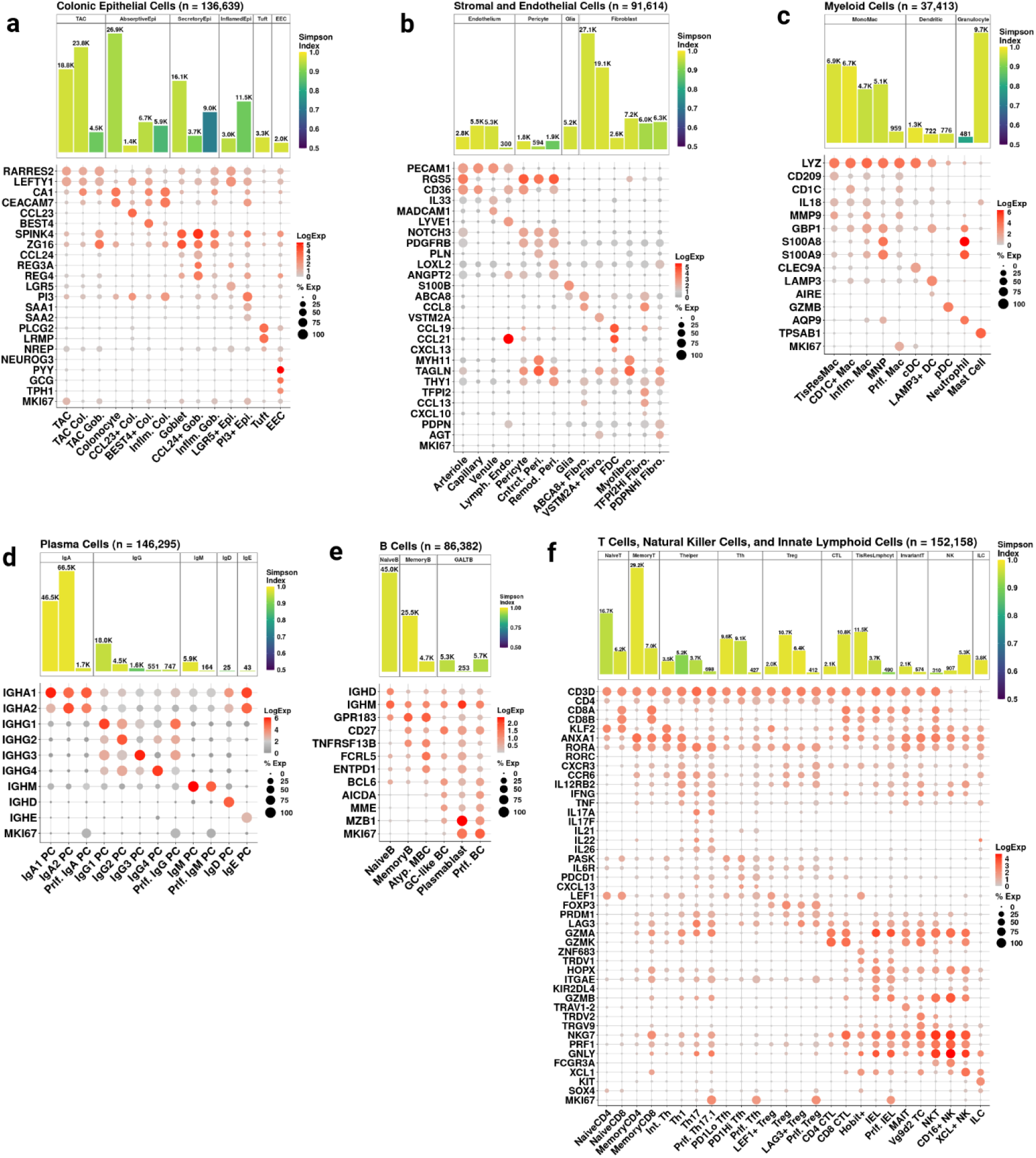
Cell state markers and biopsy diversity Cell state-defining genes within each cell group annotated in this study. Circle color denotes log-normalized expression level of gene in the annotated cell state within the six annotated major cell groups: colonic epithelium (**a**), stromal and endothelial cells (**b**), myeloid cells (**c**), plasma cells (**d**), B cells (**e**), and T, NK, and ILCs (**f**). Size of circle corresponds to percentage of cells expressing the gene in the annotated cell state. The number of cells in each cell state is denoted by the bar plot above dot plot. Fill colors correspond to Simpson’s diversity index, based on frequencies of biopsies within each cell state. Each cell state is grouped with related cell states within a cell subtype.

**Supplemental Fig. 3.**
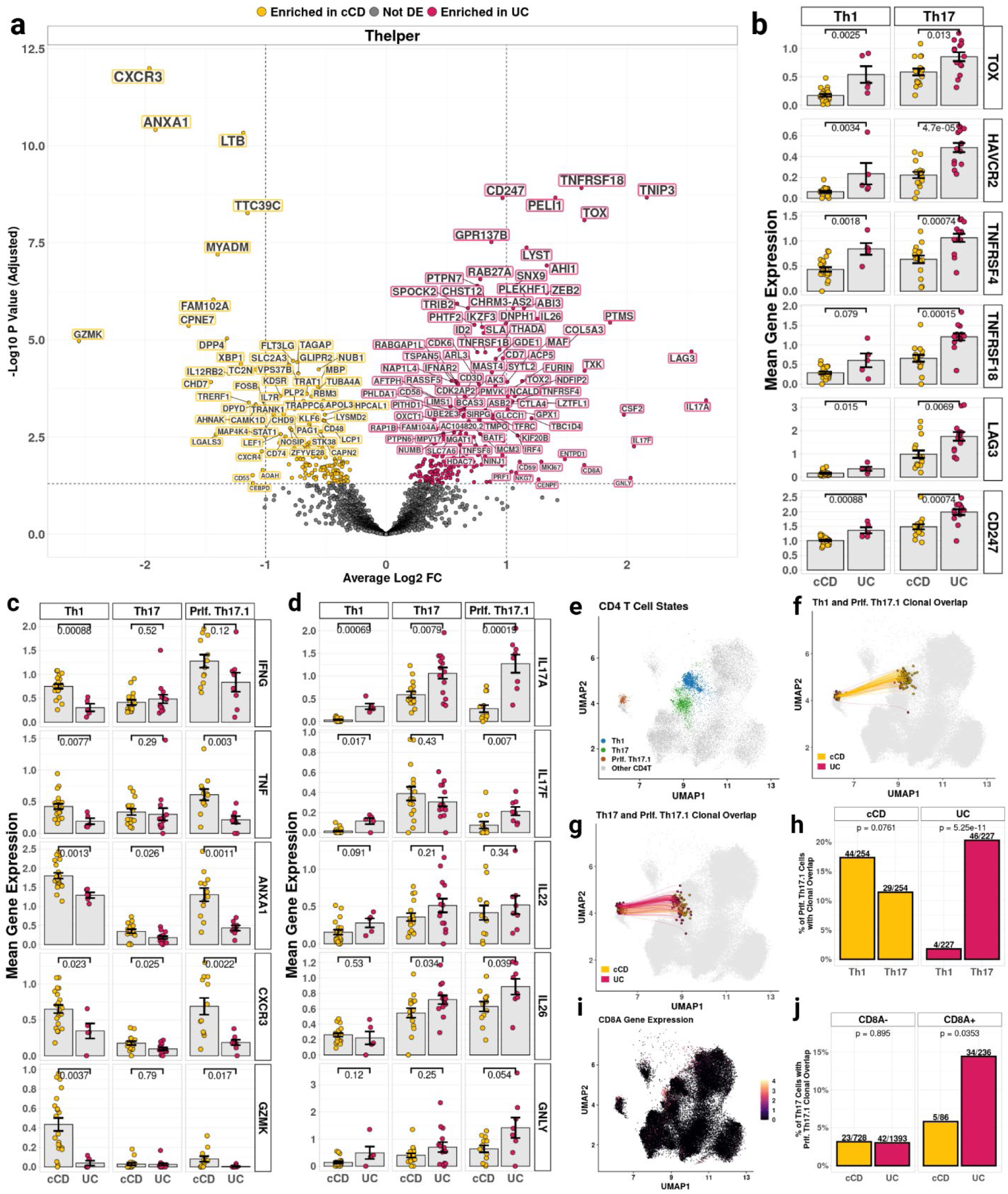
Gene expression and clonal overlap in T_H_1, T_H_17, and Proliferating T_H_17.1 cells **a**, Differential expression analysis contrasting cCD and UC gene expression within pseudobulked T_H_ cell subtype, among diagnostic biopsies with ≥ 10 T_H_ cells (cCD, n = 30; UC, n = 15). **b**, Mean log-normalized expression of T cell activation, co-stimulatory and exhaustion genes within T_H_1 and T_H_17 cells among cCD and UC biopsies with ≥ 10 T_H_1 (cCD, n = 21; UC, n = 5) or T_H_17 (cCD, n = 16; UC, n = 14). **c**, **d**, Mean log-normalized expression of T_H_1-associated (**c**) and T_H_17-associated (**d**) genes within T_H_1 and T_H_17 cells among cCD and UC biopsies with ≥ 10 T_H_1 (cCD, n = 21; UC, n = 5) or T_H_17 (cCD, n = 16; UC, n = 14). **e**, UMAP of CD4 T cells among diagnostic biopsies with T_H_1 cells (blue), T_H_17 cells (green), and proliferating T_H_17.1 cells (brown) highlighted. **f**, **g** UMAPs of shared clones between proliferating T_H_17.1 cells and T_H_1 cells (**f**) or T_H_17 cells (**g**) within cCD (yellow lines) or UC (red lines) diagnostic biopsies. Clonotypes were determined by matched paired CDR3αβ amino acid sequence. **h**, Percentage of cCD and UC proliferating T_H_17.1 cells with clonal overlap with T_H_1 or T_H_17 cells. All cells from diagnostic biopsies with at least 1 cell with paired CDR3αβ in Prlf. T_H_17.1 cell state included (cCD, n = 24; UC, n = 14). **i**, UMAP of *CD8A* expression among CD4 T cells. **j**, Percentage of cCD and UC T_H_17 cells with clonal overlap with the proliferating T_H_17 cell state, split by T_H_17 cells with or without *CD8A* expression. All cells from diagnostic biopsies with at least 1 cell with paired CDR3αβ in T_H_17 cell state included (cCD, n = 30; UC, n = 14). Error bars denote standard error of the mean. CDR, complementarity determining region.

**Supplemental Fig. 4.**
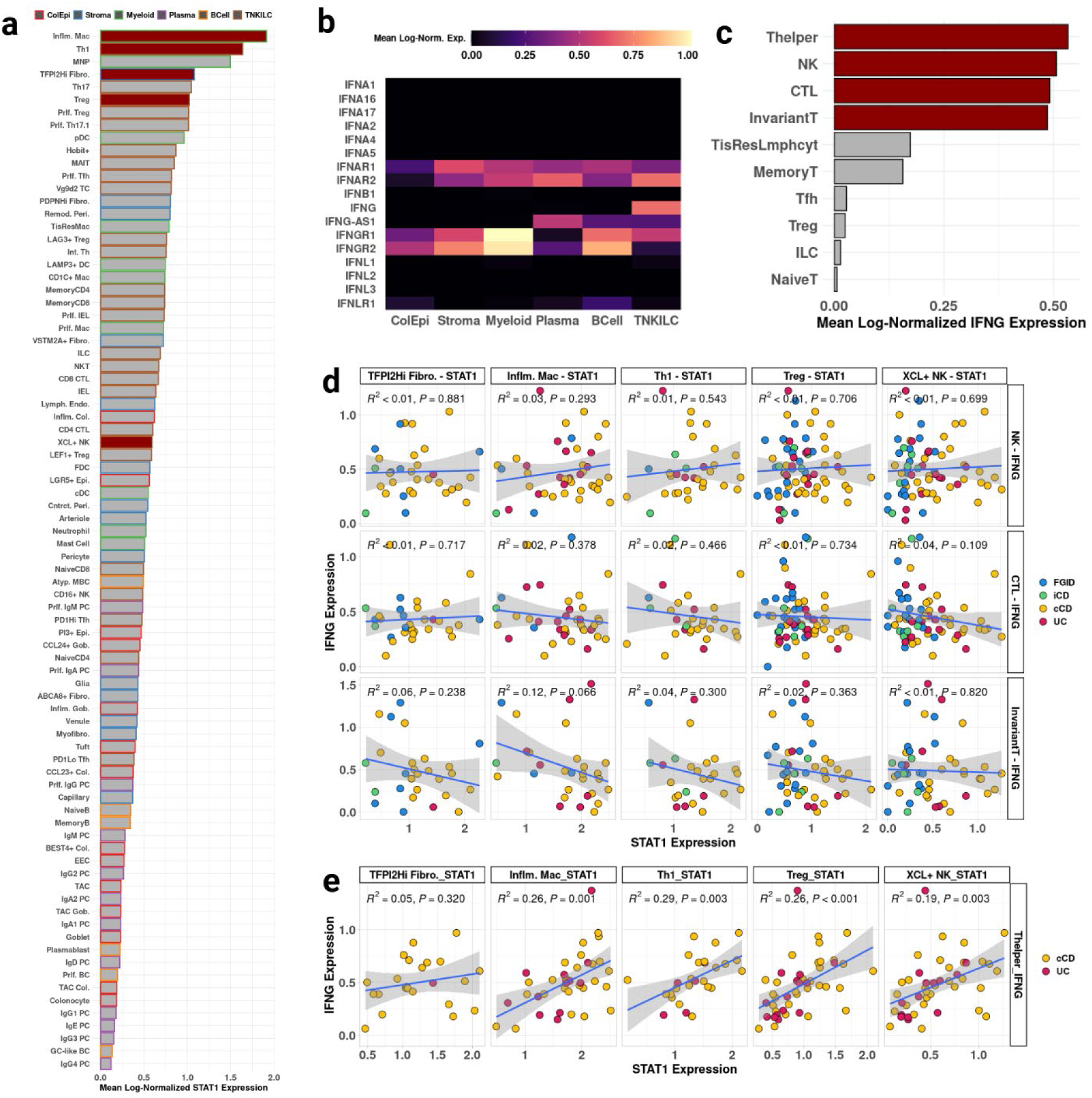
T_H_ *IFNG* expression, but not other T/NK *IFNG* expression, is correlated with *STAT1* expression in T_H_1-associated cell states **a**, Expression of *STAT1* in all cell states. Bars outlined in color corresponding to respective cell group. Bars filled red if cell state was associated with T_H_1 cell state in cell state frequency correlation analysis. **b**, Per-cell type mean log-normalized expression of type 1, type 2, and type 3 interferon cytokines and their receptors within pre-treatment biopsies. **c**, Mean log-normalized *IFNG* expression in T/NK/ILC cell subtypes pre-treatment biopsies. Bars filled red if cell subtype included in regression analyses in subfigures **d**-**e**. **d**, Relationships between NK, CTL, and InvariantT cell subtype expression of *IFNG* and T_H_1-associated cell states, modeled as simple linear regressions. **e**, Relationships between T_H_ cell subtype expression of *IFNG* and T_H_1-associated cell states in cCD and UC only, modeled as simple linear regressions. Shaded areas of regressions show 95% confidence interval.

**Supplemental Fig. 5.**
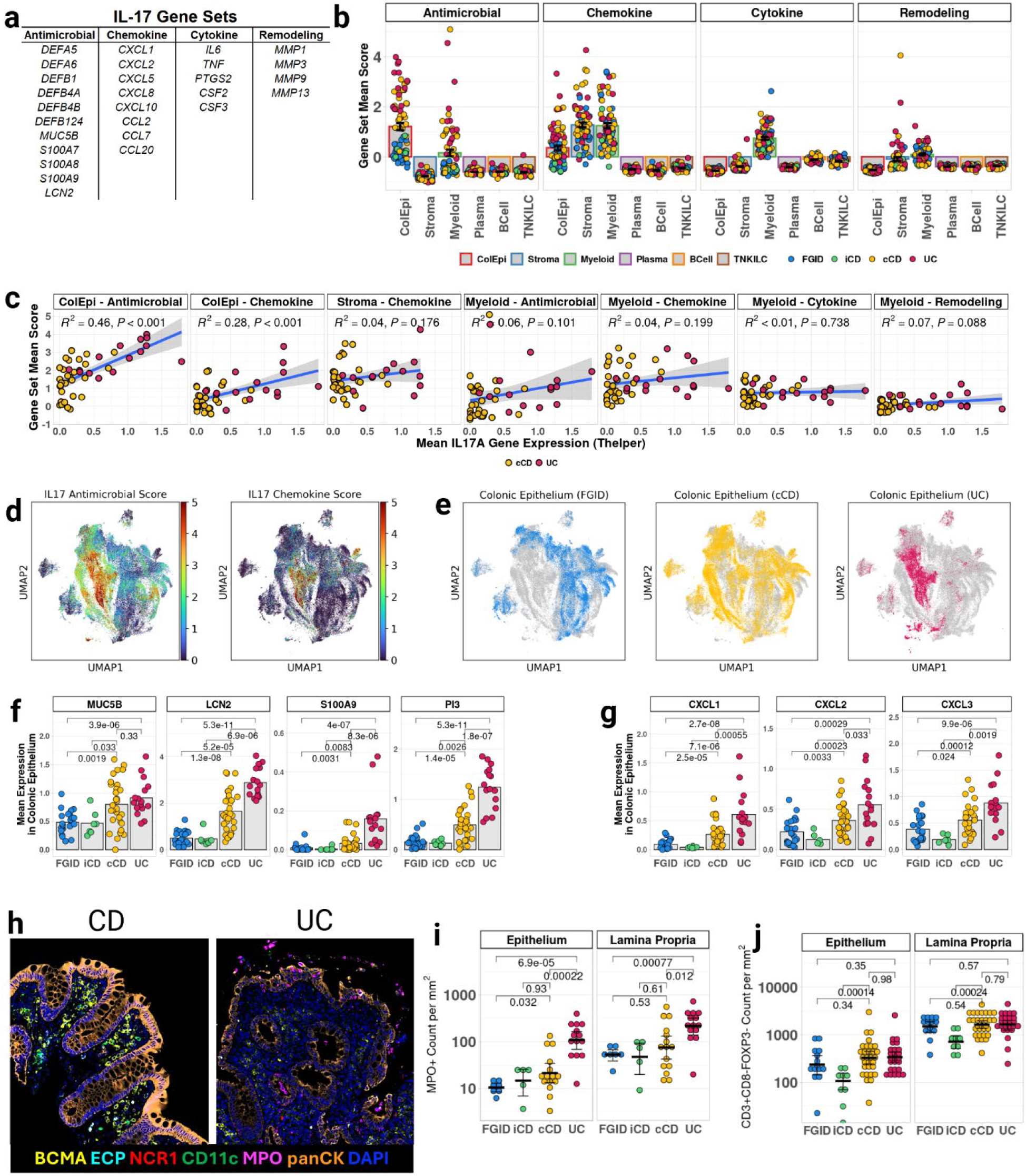
*IL17A* expression is associated with IL-17-induced antimicrobial and leukocyte recruitment genes in colonic epithelial cells **a**, Table of genes associated with IL-17A signaling in KEGG “IL-17 Signaling Pathway.” **b**, IL-17 gene set scores in diagnostic biopsies with at least 10 cells in cell type (ColEpi, n = 77; Stroma, n = 74; Myeloid, n = 74; Plasma, n = 75; BCell, n = 72; TNKILC, n = 76) **c**, Relationship between mean expression of *IL17A* in T_H_ cells and IL-17 antimicrobial and chemokine gene set scores in cell types with positive mean expression of gene set, modeled as a simple linear regression among diagnostic cCD and UC biopsies with at least 10 T_H_ cells and 10 cells in the relevant cell type (cCD, n = 30; UC, n = 14 for stromal and endothelial cells, n = 15 for all other cell types). **d**, UMAP depicting scoring of colonic epithelium for IL-17 antimicrobial (left) and chemokine (right) gene sets in diagnostic biopsies. **e**, UMAPs highlighting FGID, cCD, and UC colonic epithelial cells in diagnostic biopsies. **f**, **g**, Mean log-normalized expression of antimicrobial (**f**) or chemokine (**g**) genes in colonic epithelium, among diagnostic biopsies with ≥ 10 colonic epithelial cells (FGID, n = 23; iCD, n = 7; cCD, n = 31; UC, n = 16). **h**, Representative images from colon of Crohn’s disease and ulcerative colitis participants’ epithelium and proximal lamina propria, stained for BCMA (yellow, plasma cells), ECP (cyan, eosinophils), MPO (magenta, neutrophils), CD11c (green, dendritic cells), NCR1 (red, NK cells), DAPI (dark blue, nucleated cells), and panCK (orange, epithelium). **i**, **j**, quantification of MPO+ (**i**) and CD3+CD8-FOXP3-(**j**) cells *in situ*, separated by epithelial and lamina propria layers. All fluorescence microscopy samples taken from diagnostic biopsies (MPO: FGID, n = 7; iCD, n = 5; cCD, n = 15; UC, n = 14. CD3+CD8-FOXP3-: FGID, n = 14; iCD, n = 9; cCD, n = 30; UC, n = 25). Two-tailed Mann-Whitney U test for all pairwise comparisons. Shaded areas of regressions show 95% confidence interval.

**Supplemental Fig. 6.**
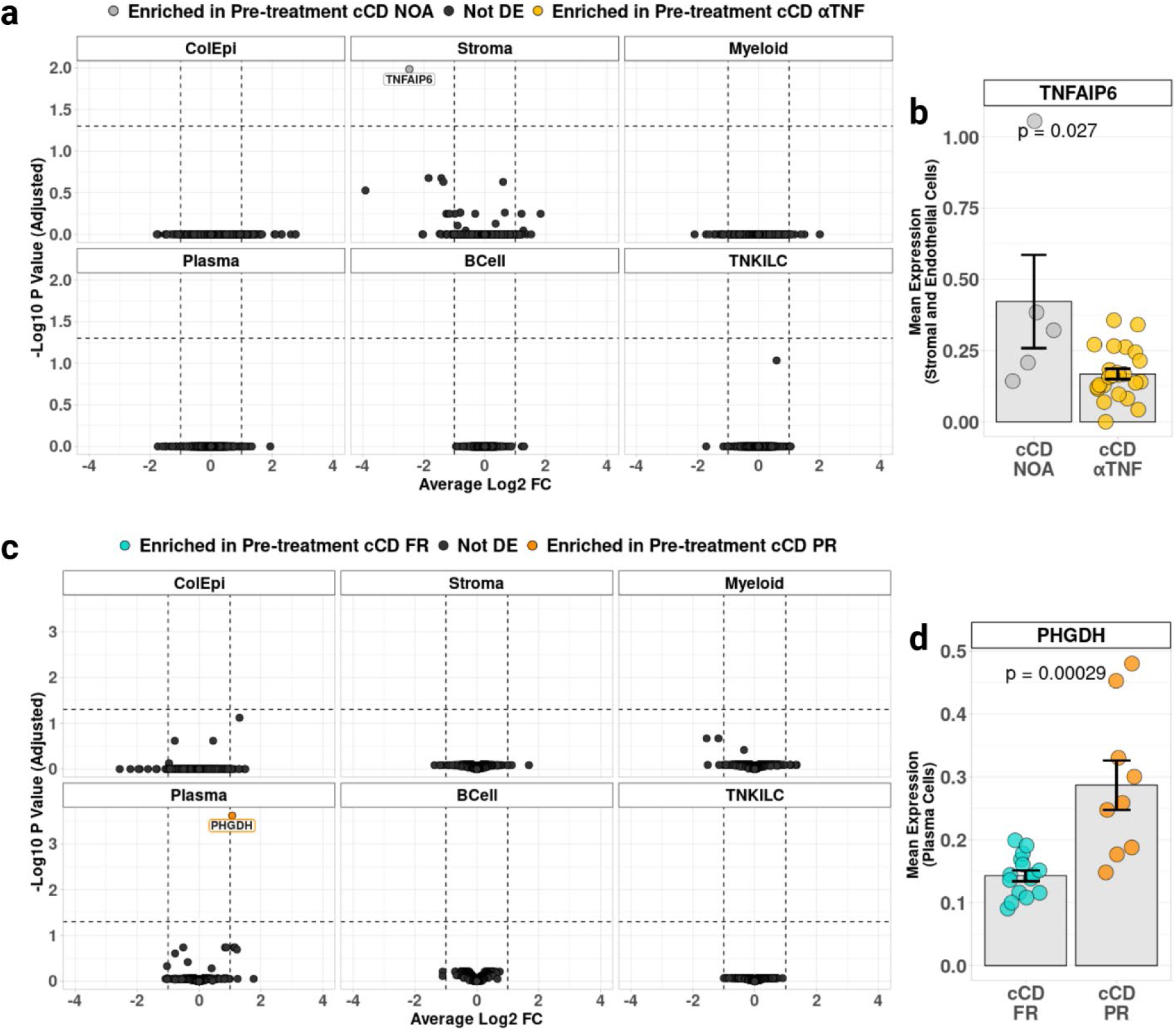
Pre-treatment transcriptomic differential expression between cCD treatment groups and anti-TNF response groups **a**, Volcano plot comparing differentially expressed genes in pre-treatment cCD NOA and anti-TNF recipients within each annotated cell type. Biopsies were included for analysis if they contained ≥ 10 cells within the cell type (NOA: n = 5 for all cell types. αTNF: stromal and endothelial cells, myeloid cells, plasma cells, n = 24; colonic epithelium, B cells, and T, NK, and ILC cells, n = 25). **b**, Mean log-normalized expression of *TNFAIP6* in stromal and endothelial cells, comparing cCD NOA and anti-TNF recipient diagnostic biopsies with ≥ 10 stromal and endothelial cells (NOA, n = 5; αTNF, n = 24). **c**, Volcano plot comparing differentially expressed genes in pre-treatment cCD FR and PR recipients within each annotated cell type. Biopsies were included for analysis if they contained ≥ 10 cells within the cell type (FR: n = 15 for all cell types. PR: stromal and endothelial cells, myeloid cells, plasma cells, n = 9; colonic epithelium, B cells, and T, NK, and ILC cells, n = 10). Two-tailed Mann-Whitney U test for all pairwise comparisons. **d**, Mean log-normalized expression of *PHGDH* in plasma cells, comparing cCD NOA and anti-TNF recipient diagnostic biopsies with ≥ 10 plasma cells (NOA, n = 5; αTNF, n = 24).

**Supplemental Fig. 7.**
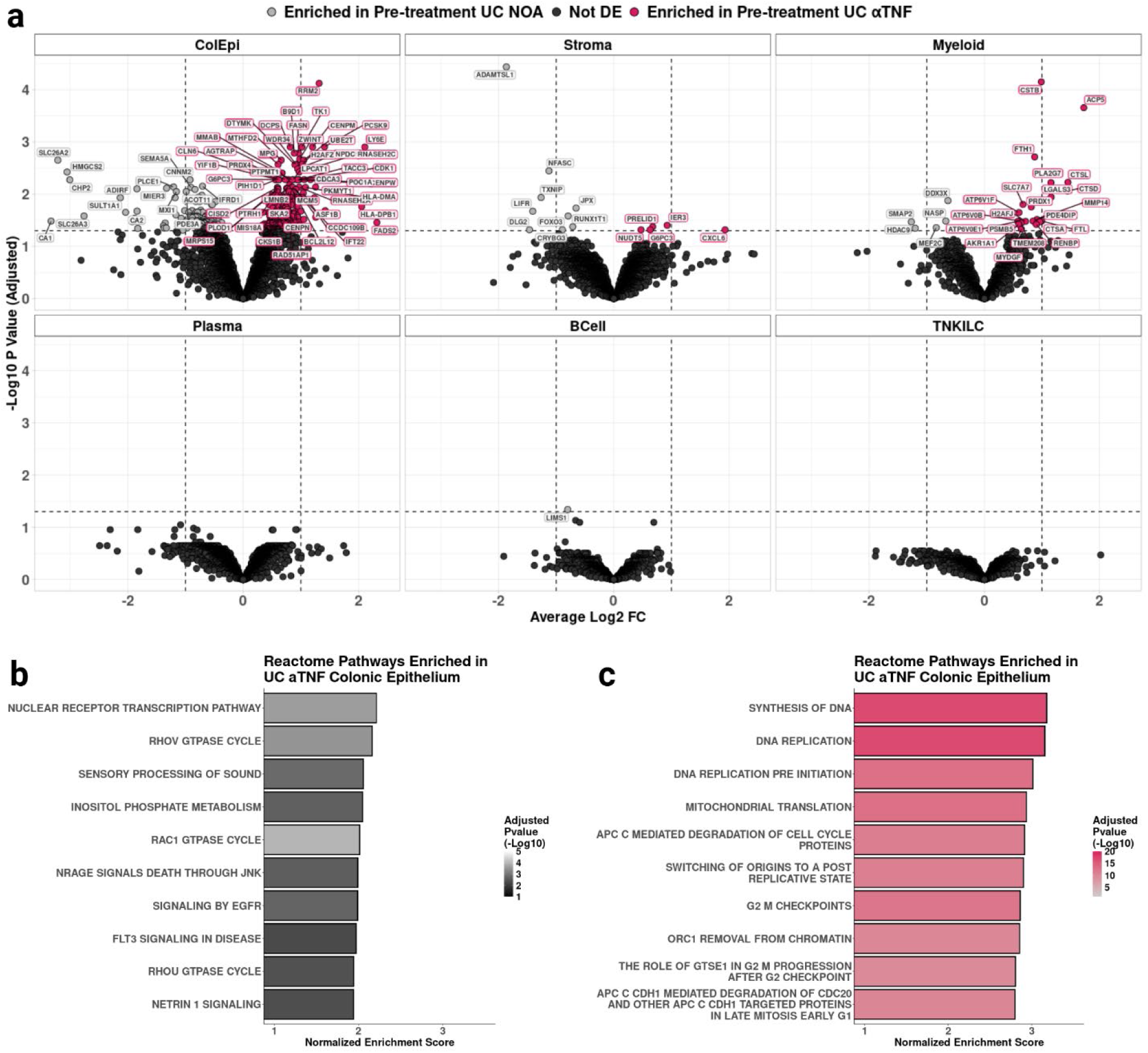
Pre-treatment transcriptomic differential expression between UC treatment groups **a**, Volcano plot comparing differentially expressed genes in UC NOA and anti-TNF recipients within each annotated cell type. Biopsies were included for analysis if they contained ≥ 10 cells within the cell type (NOA: colonic epithelium, n = 8; stromal cells, n = 6; all other cell types n = 7. αTNF: n = 8 for all cell types). **b**, **c**, Enriched terms from gene set enrichment analysis of differential expression between pre-treatment colonic epithelium from anti-TNF untreated (**b**) and treated (**c**) UC participants. Top 10 terms for each treatment group shown. Gene ranks computed using DESeq2 Wald test “stat” value (log2FC/SE(log2FC)). Ranked genes tested using Reactome 2023 v2 gene sets with 15 ≤ number of genes ≤ 100 and sorted for normalized enrichment score.

**Supplemental Fig. 8.**
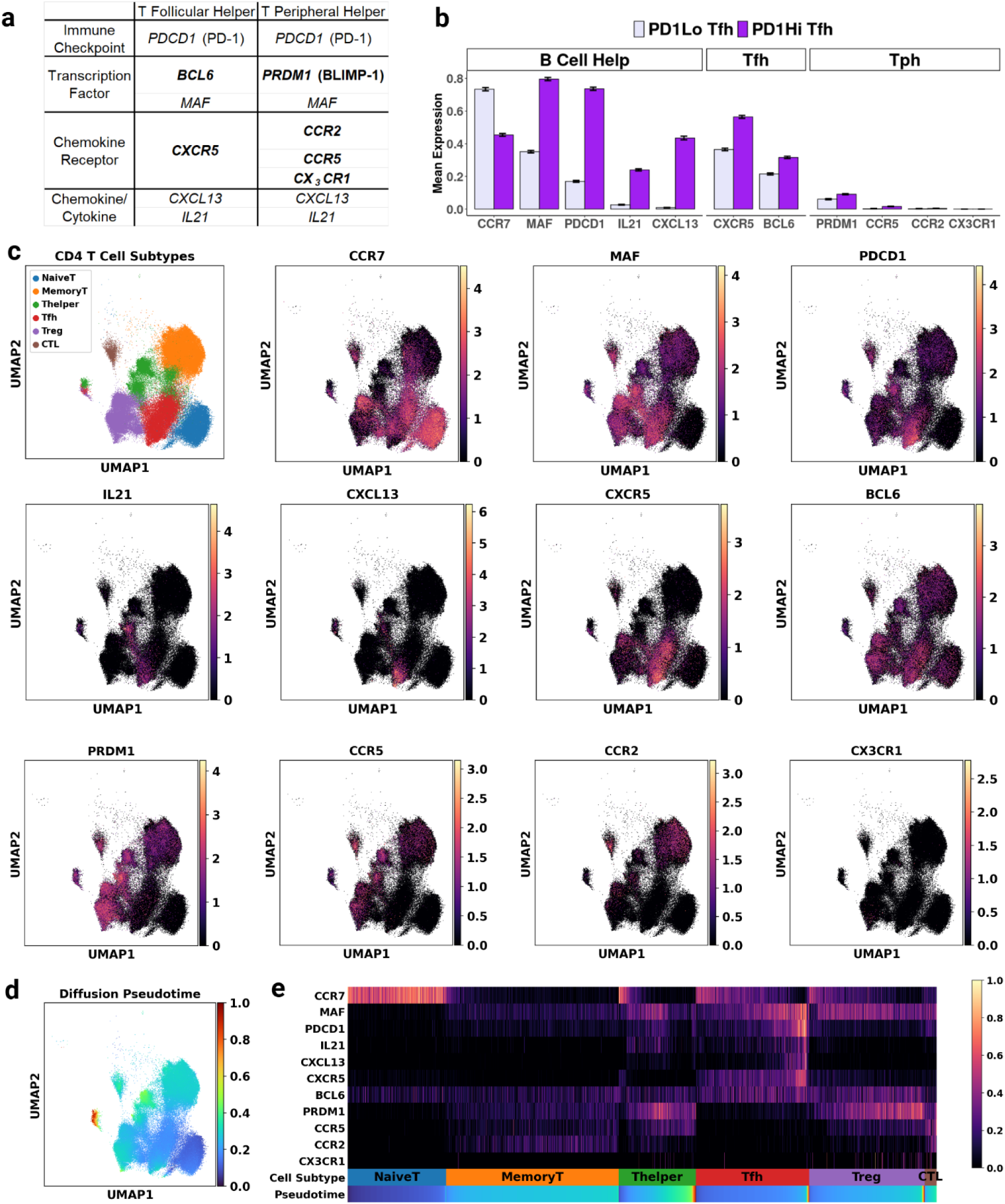
T_FH_ cells are identified without evidence of T_PH_ phenotype **a**, Table of genes canonically associated with T_FH_ and T_PH_ cells. Protein function listed in leftmost column. Bolded genes are differentiating between T_FH_ and T_PH_. **b**, Mean log-normalized expression of genes involved in B cell help, T_FH_ identity, and T_PH_ identity amongst all PD1Lo T_FH_ cells and PD1Hi T_FH_ cells. **c**, UMAPs of CD4 T cells labeled by cell state, disease group. Log-normalized expression of selected T_FH_ and T_PH_ genes overlaid on UMAP. **d**, Diffusion pseudotime UMAP of CD4 T cells. **e**, Per-gene scaled expression (0-1) of T_FH_ and T_PH_ genes among CD4 T cells, ordered by cell subtypes and pseudotime value within cell subtypes. Cells from all disease groups, treatment groups, and timepoints included for all analyses.

**Supplemental Fig. 9.**
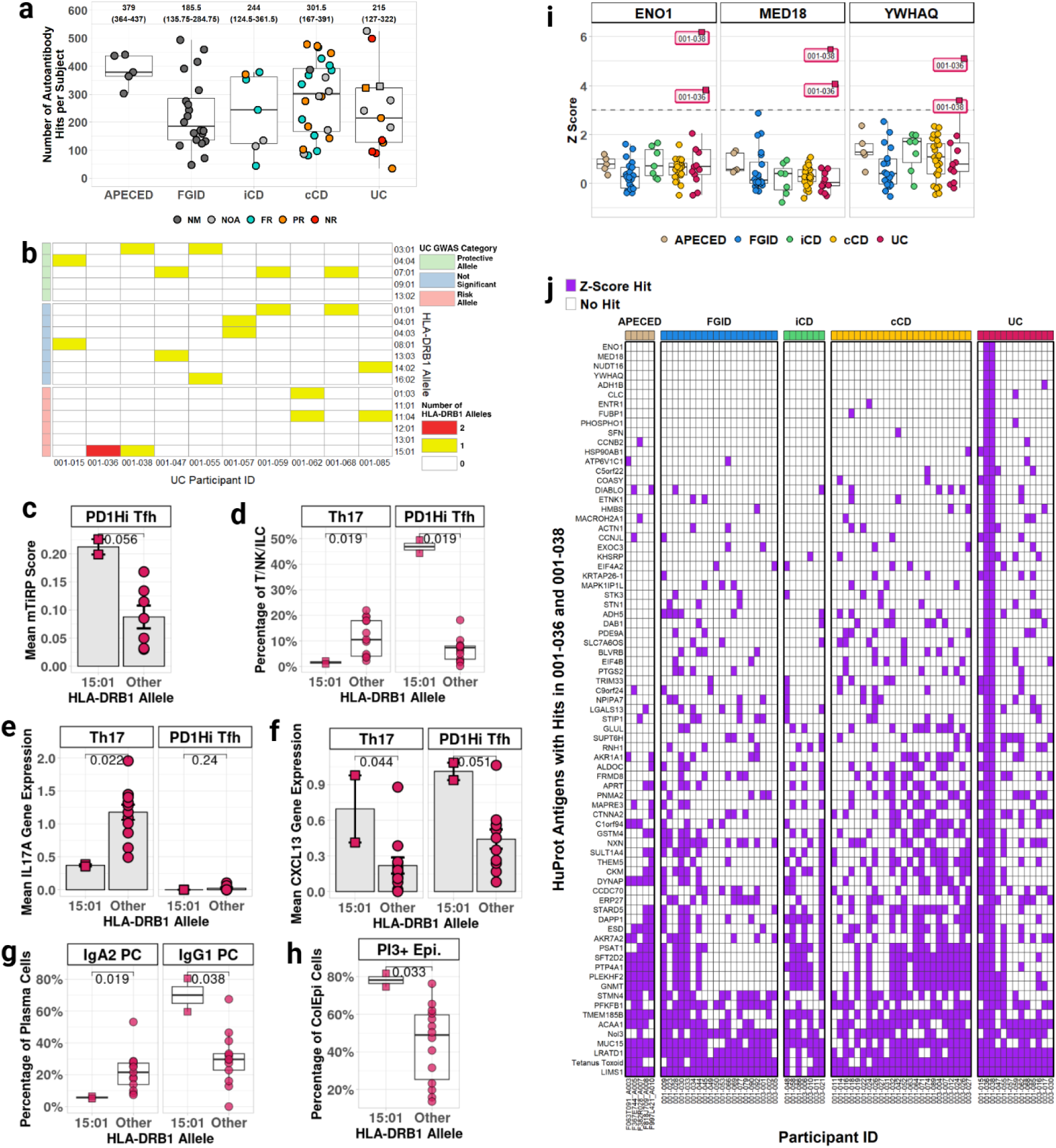
HLA-DRB1*15:01+ UC participants are skewed towards PD1Hi T_FH_-associated features and share features of autoreactivity **a**, Boxplots of number of autoantigen hits per disease group (FGID, iCD, cCD, UC, as well as 5 additional APECED samples). Median number of autoantigen hits is displayed above each group’s boxplot, followed by the range of the 25^th^ percentile and the 75^th^ percentile number of autoantigen hits. Markers are colored by anti-TNF response status. APECED, n = 5; FGID, n = 20; iCD, n = 7; cCD, n = 24; UC, n = 13. **b**, Number of HLA-DRB1 alleles in UC participants with genotype data (n = 10). **c**, Mean mTiRP score in PD1Hi T_FH_ cell state among UC diagnostic biopsies with ≥ 50 PD1Hi T_FH_ cells (with HLA-DRB1*15:01 allele, n = 2; without HLA-DRB1*15:01 allele, n = 7). **d**, Frequency of T_H_17 and PD1Hi T_FH_ cell states, relative to all T/NK/ILC, among UC diagnostic biopsies with ≥ 10 T/NK/ILCs (HLA-DRB1*15:01 allele, n = 2; without HLA-DRB1*15:01 allele, n = 13). **e**, **f**, Mean log expression of *IL17A* (**e**) and *CXCL13* (**f**) in T_H_17 and PD1Hi T_FH_ cell states, among UC diagnostic biopsies with ≥ 10 in the relevant cell state (T_H_17: with HLA-DRB1*15:01 allele, n = 2; without HLA-DRB1*15:01 allele, n = 12. PD1Hi T_FH_: with HLA-DRB1*15:01 allele, n = 2; without HLA-DRB1*15:01 allele, n = 11). **f**, **g**, Frequency of IgA2 PCs and IgG1 PCs (**g**), or PI3+ Epi. (**h**) cell states relative to their respective cell types, among UC diagnostic biopsies with ≥ 10 cells in cell type (Plasma cells: with HLA-DRB1*15:01 allele, n = 2; without HLA-DRB1*15:01 allele, n = 13. Colonic epithelium: with HLA-DRB1*15:01 allele, n = 2; without HLA-DRB1*15:01 allele, n = 14). **i**, HuProt Z-score values for 3 antigens identified as hits only in both UC participants with HLA-DRB1*15:01 allele (001-036 and 001-038). A z-score cutoff of 3 is used to denote whether an antigen is a hit within a sample (APECED, n = 5; FGID, n = 20; iCD, n = 7; cCD, n = 24; UC, n = 13). **j**, Z-score hits from HuProt proteome microarray, filtered to 77 proteins with z-score hits among both UC participants with HLA-DRB1*15:01 allele (001-036 and 001-038). Antigens are ordered by number of distinct participants with hits. Two-tailed Mann-Whitney U test for all pairwise comparisons. In subfigures with participant markers, UC participants with HLA-DRB1*15:01 allele (001-036 and 001-038) denoted by square markers instead of circle markers.

**Supplemental Fig. 10.**
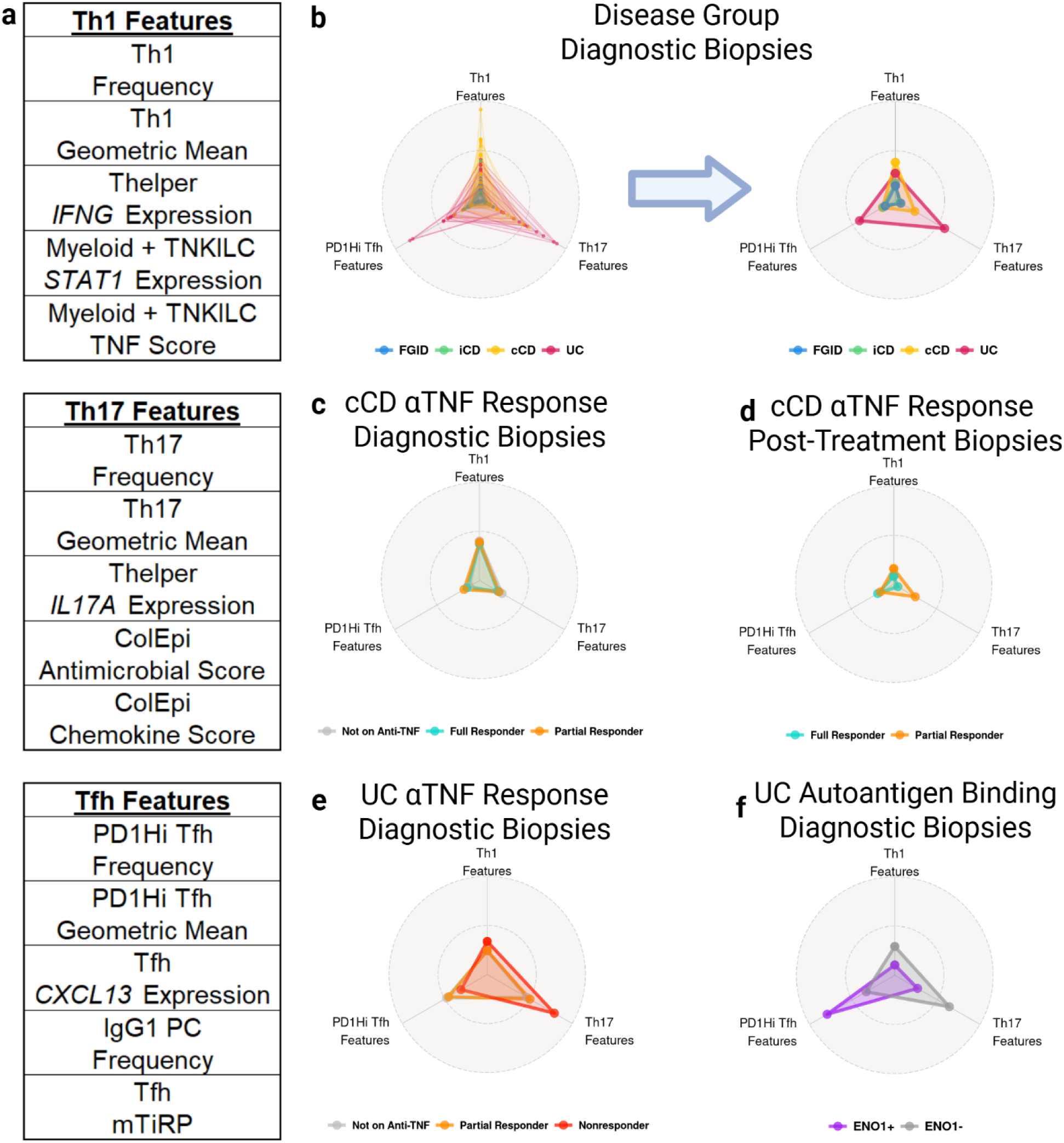
Summary of T_H_1-, T_H_17-, and PD1Hi T_FH_-associated features among disease groups, anti-TNF response categories, and antigen reactivity **a**, List of T_H_1, T_H_17, and T_FH_ features identified as distinguishing disease groups, treatment response groups, and/or autoantigen-binding groups throughout the PREDICT study. **b**, Radar plot demonstrating feature usage among diagnostic biopsies meeting all relevant cell minima and assays conducted (FGID, n = 15; iCD, n = 4; cCD, n = 19; UC, n = 10). Features were scaled between 0 and 1 (corresponding to per-feature minimum and maximum values, respectively), then averaged within each biopsy. Center of radar plot represents 0 scaled average value, outer edge represents 1 scaled average value. Radar plot shows individual diagnostic biopsies (left) or diagnostic biopsies grouped by disease (right). **c**, **d**, Radar plot comparing features among pre-treatment (**c**) and post-treatment (**d**) cCD biopsies, grouped by anti-TNF response status (Pre-treatment: NOA, n = 4; FR, n = 10; PR, n = 5. Post-treatment: FR, n = 5; PR, n = 2). **e**, Radar plot comparing features among pre-treatment UC biopsies, grouped by anti-TNF response status (NOA, n = 3; PR, n = 4; NR, n = 3). **f**, Radar plot comparing study features among pre-treatment UC biopsies, grouped by ENO1 reactivity (ENO1+, n = 2; ENO1-, n = 8).

